# Image-based deep learning for emergency electrocardiogram classification

**DOI:** 10.64898/2026.06.18.26355968

**Authors:** Felipe Meneguitti Dias, Estela Ribeiro, Natália Olivetti, Otávio Augusto Oliveira de Carvalho, Carlos Alberto Pastore, José Eduardo Krieger, Marco Antônio Gutierrez

## Abstract

Automated electrocardiogram analysis has advanced largely through digital waveforms, yet many emergency-care workflows rely on ECGs available only as printed tracings, scanned reports, PDFs or mobile photographs. We developed an image-based deep learning system for emergency ECG classification and evaluated it in InCor-EMG, an expert-adjudicated dataset of 18,519 emergency ECGs spanning 12 ECG categories, with labels from 19 cardiologists. On the held-out test set, the final ConvNeXt ensemble achieved a macro F1-score of 0.807 (95% CI, 0.788–0.825), compared with 0.820 (95% CI, 0.805–0.832) for annotating cardiologists, and higher F1-scores than Mortara Veritas^™^ in most evaluated categories. Performance was associated more strongly with inter-reader agreement than with training sample size and remained informative across scanned and photographed ECGs, with supportive performance in model-enriched temporal and heterogeneous public-image evaluations. These findings support ECG-image classification when digital waveforms are unavailable.

## 1 Introduction

The electrocardiogram (ECG) is one of the most widely used tests in emergency care. It provides rapid information about arrhythmias, acute ischemic patterns and conduction abnormalities, and timely ECG analysis can directly influence triage, reperfusion pathways and acute management [1–3].

Deep Learning (DL) has substantially advanced automated ECG analysis, with waveform-based models approaching expert-level performance across several ECG classification tasks [4, 5]. However, these models generally require access to digitally stored ECG signals. In many real-world workflows, including archives, referrals, emergency communication channels and lower-resource settings, ECGs are often available only as printed tracings, scanned reports, PDFs or mobile photographs [6–8]. These formats are readily usable by clinicians but are not directly usable by signal-based algorithms.

Image-based ECG classification offers a practical route for extending automated ECG analysis to ECGs that are stored or transmitted as images. However, evidence for this approach in expert-adjudicated emergency ECG datasets remains limited. Prior studies have often relied on synthetic renderings, single-reader labels, narrow ECG category sets or limited clinical comparators [9–12]. Emergency ECG analysis adds further complexity because clinically relevant ECG findings may be rare, visually subtle, heterogeneous and variably labeled across experts.

Here, we developed and evaluated a DL-based method for multi-label ECG image classification in emergency care. The primary benchmark was InCor-EMG, an expert-adjudicated emergency ECG-image dataset that includes 18,519 adult ECGs labeled for 12 clinically relevant ECG categories. Each ECG was reviewed independently by two cardiologists and adjudicated by a third cardiologist when required. Model development was supported by two additional ECG-image datasets used as pre-training before final training on InCor-EMG: ECGIMGSin, a synthetic ECG-image corpus generated from public waveforms, and InCor-AMB, a real ambulatory ECG-image dataset from *Instituto do Coração do Hospital das Clínicas da Universidade de S*ão *Paulo* (InCor-HCFMUSP).

We evaluated the final model on a held-out emergency test set and compared its performance with annotating cardiologists and with Mortara Veritas^™^ commercial ECG software. We also assessed performance under common image-domain shifts using scanned and photographed ECGs and in two complementary evaluations: a model-enriched future emergency ECG subset from a different device and layout, and heterogeneous public ECG images. Finally, we examined whether between-class variation in model performance was more closely associated with training sample size or with visual ambiguity, using inter-reader agreement as an interpretable measure of label consistency.

## 2 Results

### 2.1 InCor-EMG cohort and expert adjudication

InCor-EMG included 18,519 adult emergency ECG examinations from 14,402 unique patients, represented as images with dimensions of 3122 by 1671 pixels and three color channels. A representative full ECG image in this format is shown in Supplementary Figure 1. ECGs were labeled under a multi-label taxonomy comprising 12 emergency-relevant ECG categories: Normal, Atrial Fibrillation (AF), Atrial Tachycardia or Atrial Flutter (AT/Flutter), Paroxysmal Supraventricular Tachycardia or Sinus Tachycardia (PSVT/ST), Right Bundle Branch Block (RBBB), Left Bundle Branch Block (LBBB), Second-degree, Third-degree or Advanced Atrioventricular Block or Sinus Bradycardia (AVB 2°/3°/advanced/SB), Premature beat, Pacemaker rhythm (PM), ST elevation, Injury current and Ventricular Tachycardia or Ventricular Fibrillation (VT/VF).

Representative ECG patterns from leads II, V1 and V5 for each modeled ECG category are shown in Supplementary Figure 2. ECGs without any target-class finding were assigned to Other. Cohort characteristics and class frequencies are shown in Table 1. The complete taxonomy and annotation criteria are described in the Methods.

**Table 1.**
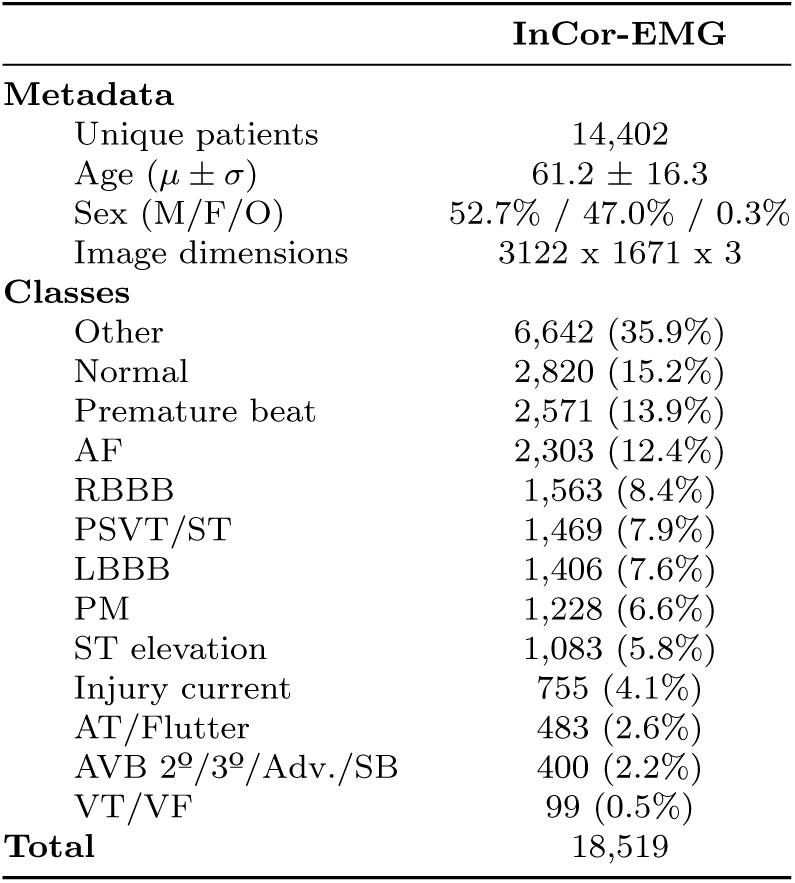
Summary of the InCor-EMG analytical cohort. Percentages for ECG categories were computed per ECG under the multi-label formulation.

| InCor-EMG |  |
| --- | --- |
| **Metadata** |  |
| Unique patients | 14,402 |
| Age ($\mu \pm \sigma$) | 61.2 ± 16.3 |
| Sex (M/F/O) | 52.7% / 47.0% / 0.3% |
| Image dimensions | 3122 x 1671 x 3 |
| **Classes** |  |
| Other | 6,642 (35.9%) |
| Normal | 2,820 (15.2%) |
| Premature beat | 2,571 (13.9%) |
| AF | 2,303 (12.4%) |
| RBBB | 1,563 (8.4%) |
| PSVT/ST | 1,469 (7.9%) |
| LBBB | 1,406 (7.6%) |
| PM | 1,228 (6.6%) |
| ST elevation | 1,083 (5.8%) |
| Injury current | 755 (4.1%) |
| AT/Flutter | 483 (2.6%) |
| AVB 2<sup>o</sup>/3<sup>o</sup>/Adv./SB | 400 (2.2%) |
| VT/VF | 99 (0.5%) |
| **Total** | 18,519 |

To obtain emergency reference labels, InCor-EMG was divided into 20 annotation batches. Each ECG was independently reviewed by two cardiologists using a secure web-based platform, and disagreements in modeled ECG categories were adjudicated by a third cardiologist. Nineteen cardiologists participated in the annotation process. The annotation and adjudication workflow is shown in Supplementary Figure 4.

Inter-reader agreement varied widely across ECG categories (Table 2). Pacemaker rhythm showed almost perfect agreement (*κ* = 0.855, 95% CI, 0.821–0.883), whereas ST elevation showed only fair agreement (*κ* = 0.313, 95% CI, 0.263–0.363). Overall, rhythm and conduction categories tended to show higher agreement than ST-segment or more heterogeneous categories.

**Table 2.** Cohen’s kappa by ECG category across the 20 InCor-EMG annotation batches.

| Class | $\kappa$ (95% CI) | Agreement |
| --- | --- | --- |
| PM | 0.855 (0.821–0.883) | Almost perfect |
| Premature beat | 0.802 (0.782–0.822) | Substantial |
| AF | 0.791 (0.758–0.823) | Substantial |
| RBBB | 0.772 (0.738–0.805) | Substantial |
| LBBB | 0.647 (0.608–0.688) | Substantial |
| AVB 2<sup>o</sup>/3<sup>o</sup>/Adv./SB | 0.609 (0.552–0.661) | Substantial |
| PSVT/ST | 0.574 (0.498–0.642) | Moderate |
| Injury current | 0.560 (0.506–0.610) | Moderate |
| Normal | 0.520 (0.446–0.592) | Moderate |
| VT/VF | 0.478 (0.353–0.586) | Moderate |
| AT/Flutter | 0.440 (0.361–0.522) | Moderate |
| Other | 0.344 (0.299–0.387) | Fair |
| ST elevation | 0.313 (0.263–0.363) | Fair |

The physician annotation process was also characterized at the annotator level. Figure 1 summarizes physician-level F1-score against the adjudicated reference standard, years of ECG reading experience and mean annotation time per ECG. These reader-level summaries complement the class-level agreement analysis by describing the human annotation process underlying the adjudicated reference standard.

**Fig. 1.**
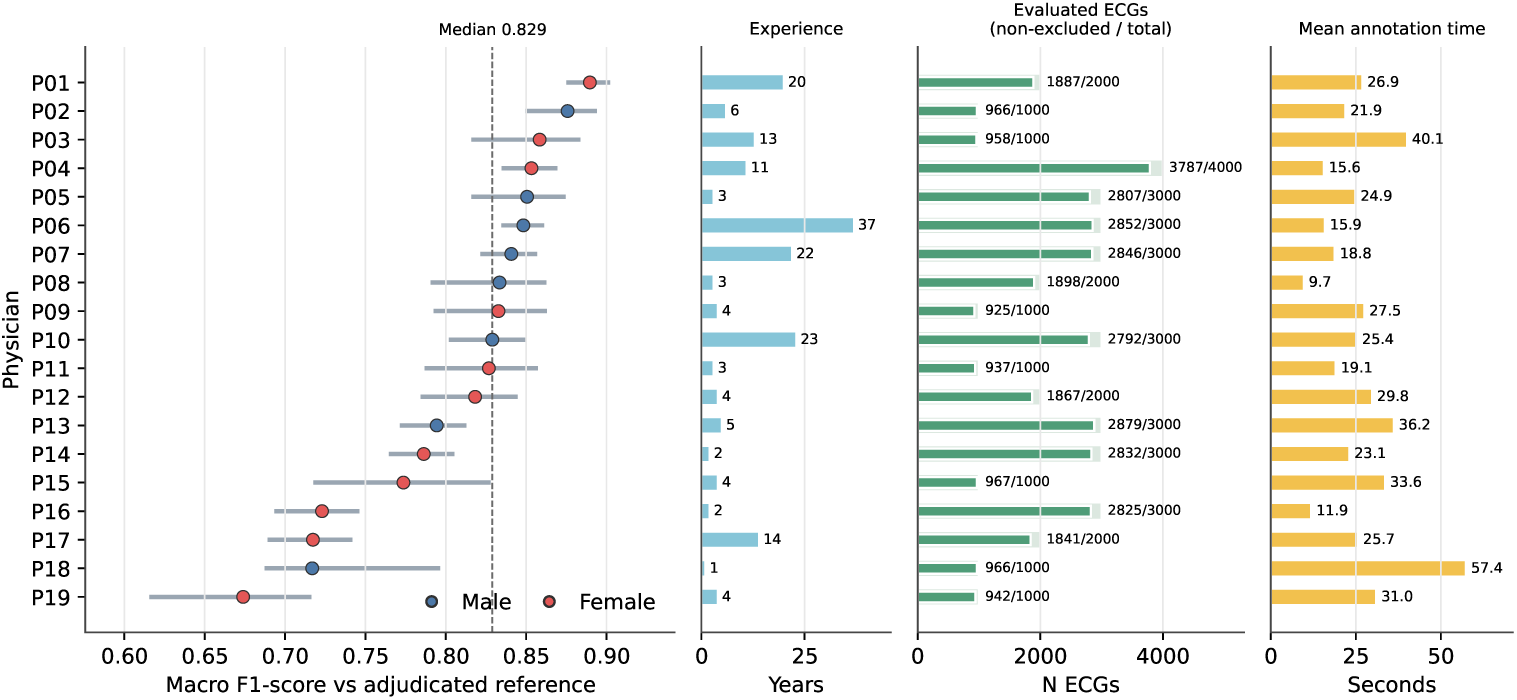
Physician annotation profile in the InCor-EMG dataset. The panels show physician-level F1-score against the adjudicated reference standard, ECG reading experience, sex distribution and mean annotation time per ECG among participating cardiologists.

A patient-level split reserved 20% of InCor-EMG as a held-out internal test set. The remaining 80% was used for model development through 10-fold cross-validation. Patient-level separation was enforced across all splits to avoid leakage from repeated ECGs from the same patient. The complete split structure is shown in Supplementary Figure 5.

### 2.2 Model development and held-out emergency performance

Model development used ECGIMGSin and InCor-AMB before final training on InCor-EMG. ECGIMGSin contained more than two million synthetic ECG images generated from public ECG waveforms and was used for ECG-domain pretraining. InCor-AMB was a previously described ambulatory ECG-image dataset from InCor-HCFMUSP with 100,450 real ECG images [12], providing additional exposure to institutional ECG-image morphology. Dataset construction details are reported in the Methods, and Supplementary Table 2 summarizes these datasets.

Within the InCor-EMG development set, ConvNeXt achieved the highest mean class-wise F1-score among five candidate image-classification backbones, with a mean F1-score of 0.737 across 10 cross-validation folds. Subsequent optimization improved the cross-validated mean class-wise F1-score to 0.777 using data augmentation, ImageNet plus ECG-domain pretraining, a custom loss function and high-resolution retraining. Architecture-selection and ablation results are provided in Supplementary Tables 3 and 4.

The final model was an ensemble of the 10 optimized ConvNeXt models trained during cross-validation on the development set. On the held-out InCor-EMG test set, the ensemble achieved a macro AUROC of 0.984 (95% CI, 0.981–0.986), macro AUPRC of 0.865 (95% CI, 0.847–0.882) and macro F1-score of 0.807 (95% CI, 0.788–0.825). Full class-wise internal test metrics are provided in Supplementary Table 5.

### 2.3 Comparison with cardiologists and Mortara Veritas^™^

The model was compared with the 19 cardiologists who annotated the dataset, using the adjudicated reference standard as the target. Because the adjudicated reference standard was derived from the same annotation process, cardiologist performance should be interpreted as an annotator-consistency benchmark rather than as an independent reader-study estimate. The mean cardiologist F1-score on the held-out test set was 0.820 (95% CI, 0.805–0.832), compared with 0.807 (95% CI, 0.788–0.825) for the model. The difference in macro F1-score was not statistically significant in paired resampling analysis. At the class level, statistically significant differences favored cardiologists for ST elevation and pacemaker rhythm, whereas the model achieved higher point estimates for selected rhythm, conduction and emergency-actionable categories, including atrial fibrillation, ventricular tachycardia/fibrillation, injury current and right bundle branch block (Figure 2 and Supplementary Table 6).

**Fig. 2.**
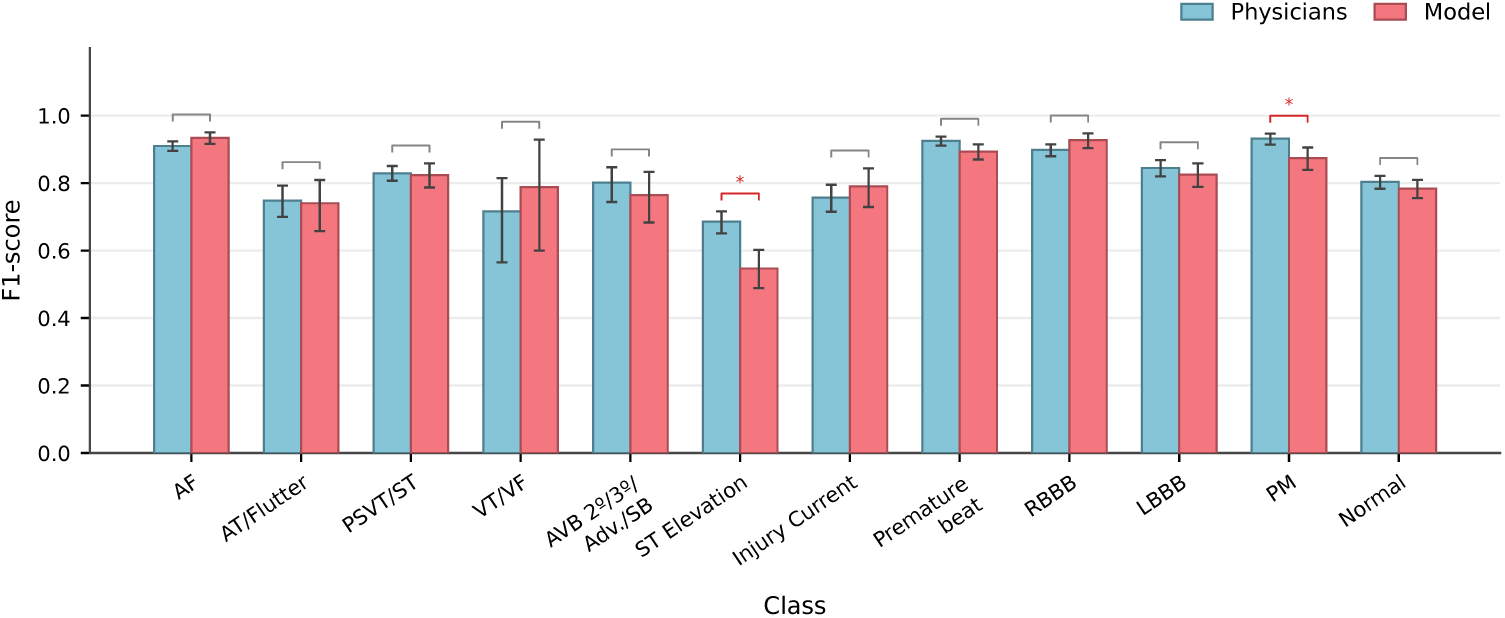
Class-wise comparison of F1-score between the proposed model and annotating cardiologists on the held-out InCor-EMG test set. Bars represent mean F1-score and error bars indicate 95% confidence intervals. Asterisks indicate statistically significant differences after Holm adjustment.

Mortara Veritas^™^ software outputs were available for 3,620 ECGs from the held-out InCor-EMG test set. After cardiologist mapping of Mortara textual statements to the modeled ECG categories, the proposed model achieved higher point F1-scores than mapped Mortara Veritas outputs in all evaluated categories, with statistically significant differences in all categories except pacemaker rhythm. This comparison should be interpreted as a pragmatic benchmark against commercial ECG statements mapped to the study taxonomy, rather than as a direct head-to-head comparison between systems natively configured for the same multi-label output space. The largest gains occurred in several acute ECG categories, including atrial tachycardia/flutter, supraventricular tachycardia/sinus tachycardia, ventricular tachycardia/fibrillation, atrioventricular block/bradycardia, ST elevation and injury current. Pacemaker rhythm was the only category without a statistically significant difference (Figure 3 and Supplementary Table 7).

**Fig. 3.**
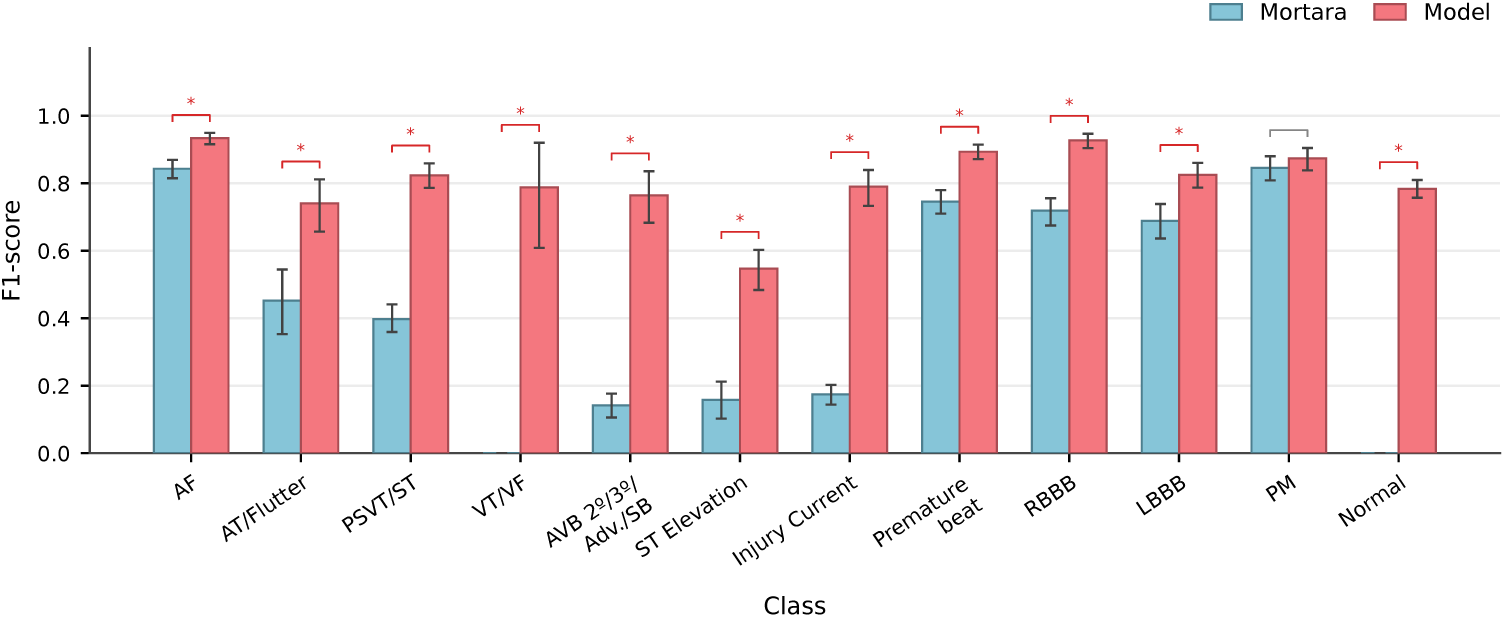
Class-wise comparison of F1-score between Mortara Veritas^™^ and the proposed model on the subset of held-out InCor-EMG test ECGs with available commercial software outputs. Bars represent mean F1-score and error bars indicate 95% confidence intervals. Asterisks indicate statistically significant differences after Holm adjustment.

### 2.4 Association between reader agreement and model performance

Training sample size is often assumed to be a major determinant of DL performance. We therefore tested whether class-level performance was more closely associated with the number of positive training examples or with inter-reader agreement. Across the 12 modeled ECG categories, model performance was more consistently associated with Cohen’s *κ* than with the log10-transformed number of positive training examples (Figure 4).

**Fig. 4.**
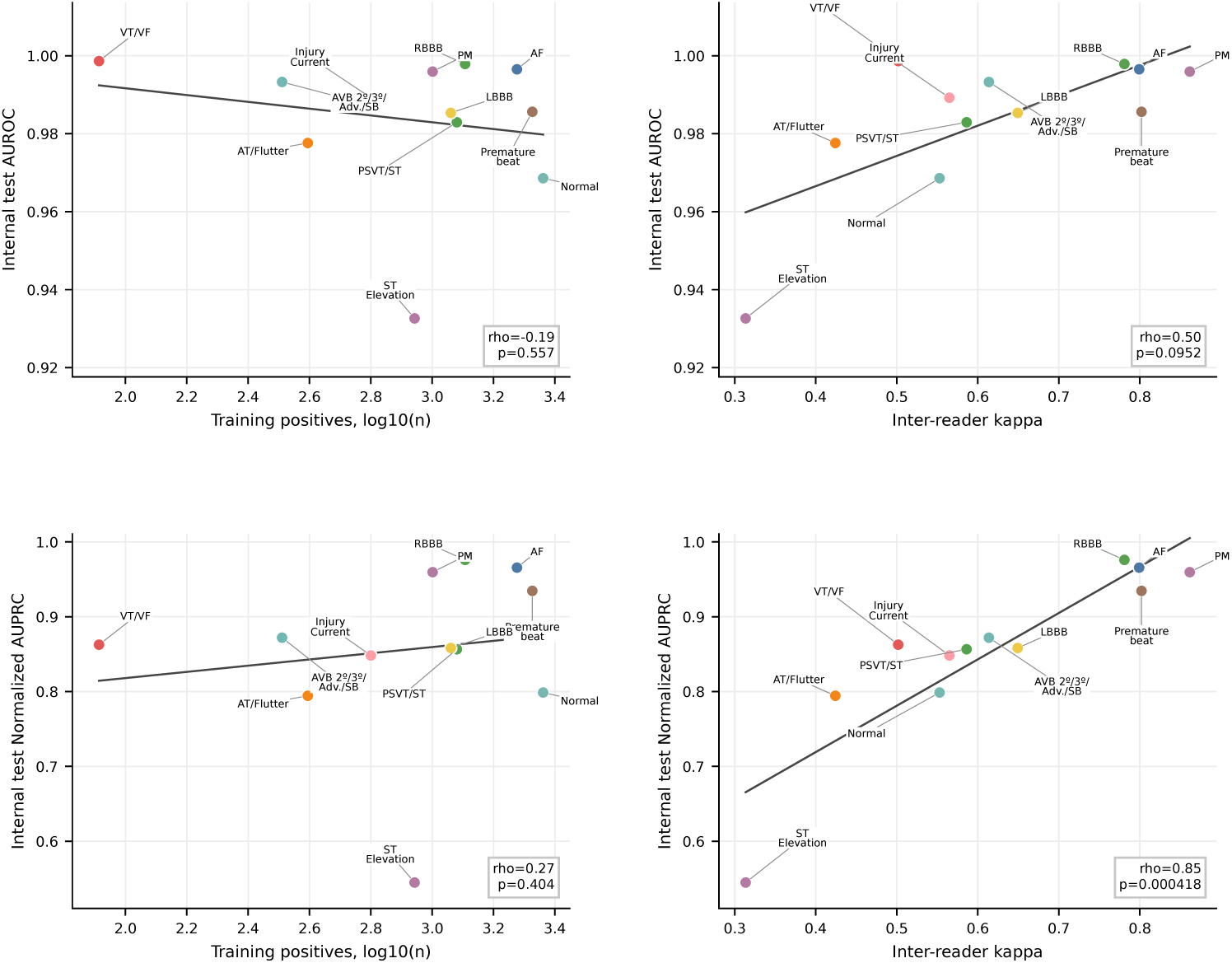
Class-level associations between model performance, inter-reader agreement and positive training examples. Performance was more strongly associated with Cohen’s *κ* than with the log10-transformed number of positive training examples, suggesting that lower inter-reader agreement, likely reflecting greater visual and labeling ambiguity, was more strongly associated with lower class-wise model performance than training sample size.

This association persisted in multivariable class-level models that included both standardized inter-reader *κ* and log10-transformed positive training examples. Inter-reader agreement showed the stronger descriptive association with model performance across F1-score, AUROC and normalized AUPRC, whereas the number of positive training examples did not show a consistent positive association in this class-level analysis. Full descriptive regression coefficients, in-sample *R*^2^ and leave-one-class-out *R*^2^ values are provided in Supplementary Table 8. Because this analysis included only 12 ECG categories, the multivariable coefficients should be interpreted as unstable and hypothesis-generating rather than as evidence of independent determinants of model performance. Together, these class-level analyses suggest that lower inter-reader agreement, likely reflecting greater visual and labeling ambiguity, was associated with lower model performance.

We next asked whether the same pattern could be observed within each ECG category. For this analysis, held-out test ECGs were stratified according to whether the two primary cardiologist readers agreed or disagreed before adjudication. Across categories, the model showed stronger discrimination among ECGs with reader agreement and lower discrimination among ECGs that required adjudication (Figure 5). Thus, the categories with lower reader agreement were not only harder overall; within categories, the cases that generated disagreement between readers were also the cases in which model ranking performance was reduced. This finding supports the interpretation that a meaningful component of model error concentrated near ECG patterns that also generated disagreement among human readers.

**Fig. 5.**
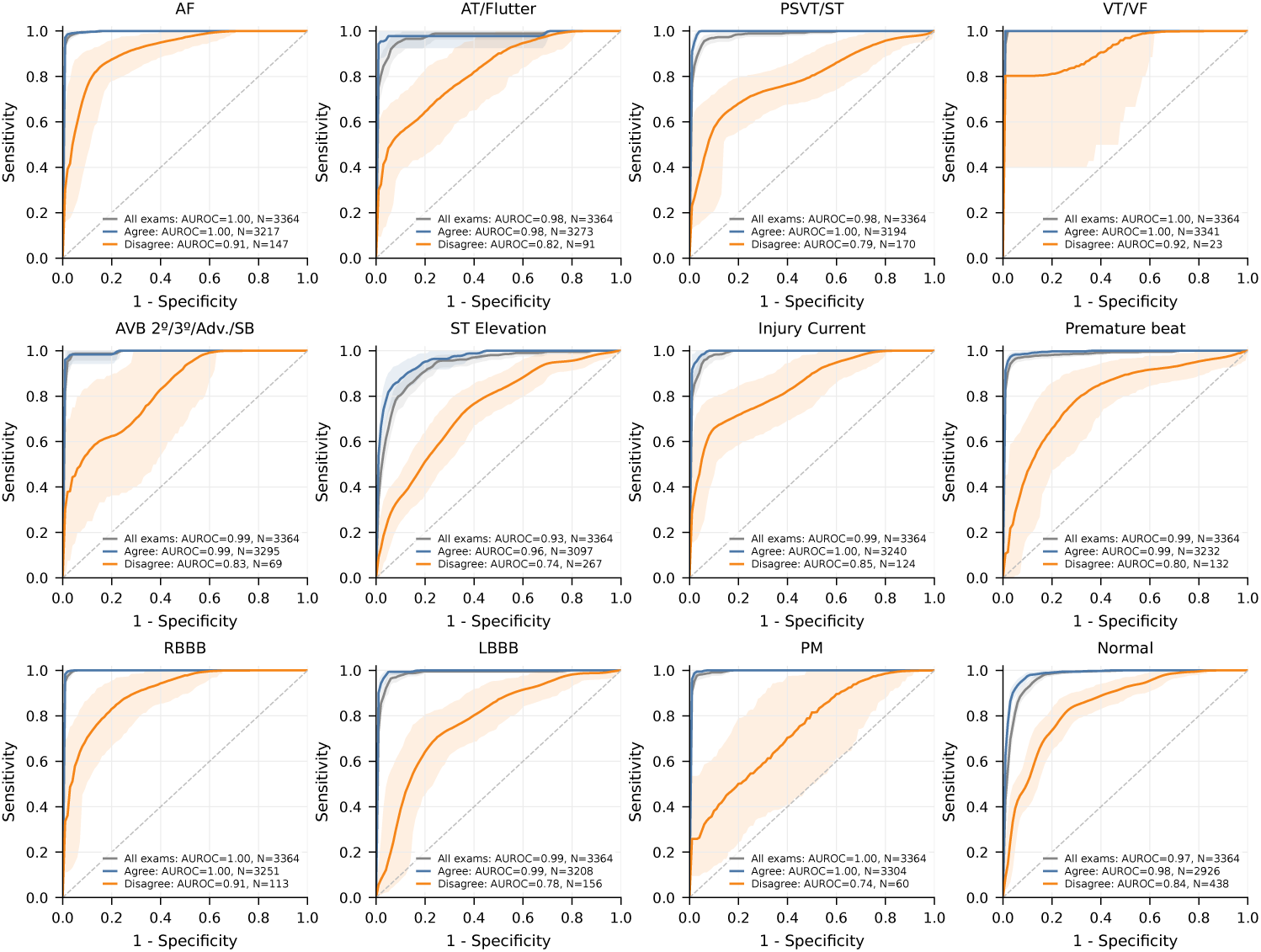
Class-wise receiver operating characteristic curves stratified by primary-reader agreement. Curves are shown for all test ECGs, ECGs in which the two primary cardiologist readers agreed and ECGs in which they disagreed before adjudication. Model discrimination was consistently higher in reader-agreement cases and lower in reader-disagreement cases, indicating that reduced model performance concentrated near visually ambiguous ECG patterns.

### 2.5 Performance across acquisition-shift, model-enriched temporal and external image evaluations

The acquisition-shift analysis used a 120-ECG subset constructed from the held-out InCor-EMG test set by randomly selecting 10 examples for each modeled ECG category. Each selected ECG was evaluated under three acquisition conditions: original digital image, scanned printed ECG and smartphone photograph, with examples shown in Supplementary Figure 10. Macro F1-score was 0.823 (95% CI, 0.763– 0.865) for original digital images, 0.752 (95% CI, 0.670–0.802) for scanned ECGs and 0.774 (95% CI, 0.702–0.823) for photographed ECGs. Macro AUROC was 0.981 (95% CI, 0.970–0.990) in original images, 0.974 (95% CI, 0.955–0.986) in scanned images and 0.974 (95% CI, 0.956–0.987) in photographed images. No class-wise F1-score or AUROC decrease remained statistically significant after Holm adjustment (Supplementary Tables 9, 10 and 11).

We next performed a model-enriched temporal emergency evaluation using future ECGs acquired after the InCor-EMG study period. The source images were acquired in 2023–2024 from a GE-MUSE system, whereas the InCor-EMG training and internal test ECGs were exported from Mortara ELI 250c devices. This evaluation therefore introduced a combined temporal, device and layout shift. A representative ECG image from this evaluation is shown in Supplementary Figure 11. The temporal separation between the retrospective InCor-EMG cohort and the future GE-MUSE emergency ECGs is shown in Figure 6.

**Fig. 6.**
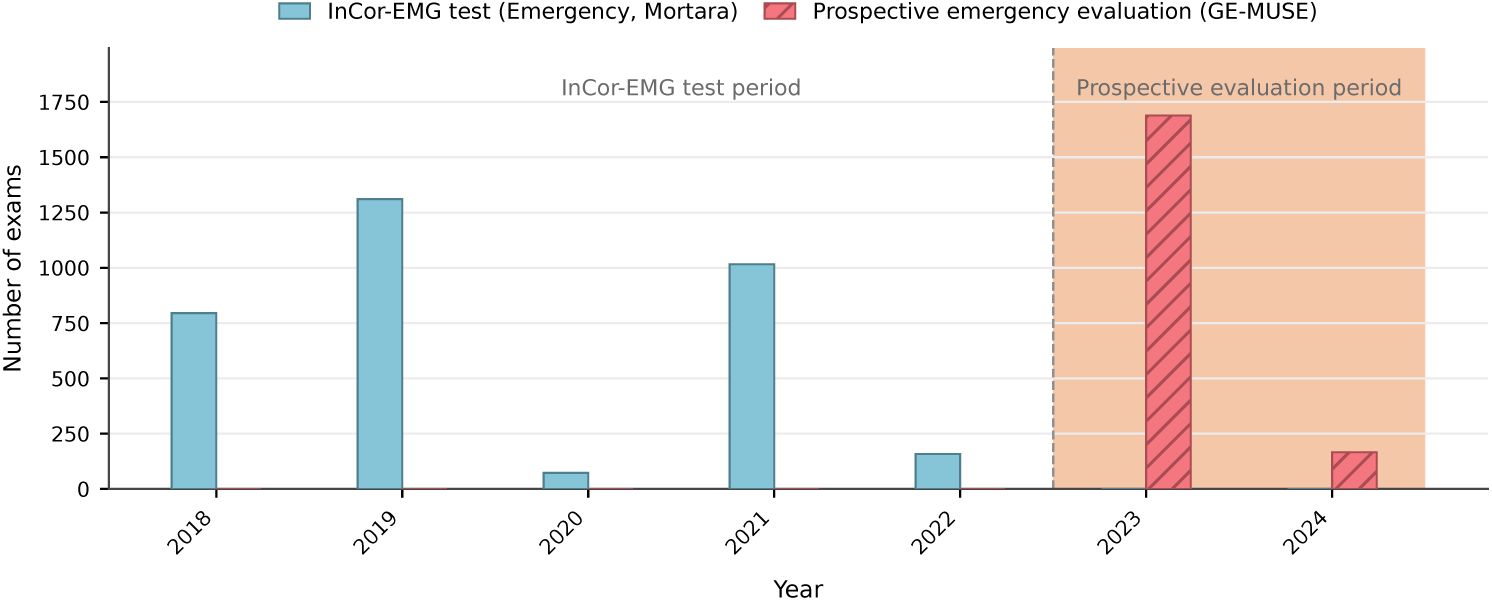
Distribution of emergency ECG examinations by acquisition year. The figure shows the temporal separation between the retrospective InCor-EMG cohort used for model development and internal testing and the future GE-MUSE emergency ECGs used for the model-enriched temporal evaluation.

This cohort was not intended to represent an unselected prospective emergency validation cohort. Future ECGs were first processed by the previously described Dias et al. model [12], which classifies ECG images as AF, Normal or Other. We selected ECGs classified as Other and negative for both AF and Normal by that earlier model, without using the proposed model or its predictions for case selection. This design enriched the temporal cohort for non-normal, non-AF ECGs requiring finer emergency-category classification. The selected ECGs were then labeled by a single cardiologist using the same ECG category criteria applied in InCor-EMG. After exclusion of technically invalid examinations, 1,855 emergency ECG images were included.

Because the cohort was enriched through a previous image-based model and labeled by a single cardiologist rather than independently reviewed and adjudicated, this analysis was treated as a model-enriched temporal stress test rather than as an unselected or expert-adjudicated validation cohort. The final ensemble achieved a macro AUROC of 0.962 (95% CI, 0.956–0.968), macro AUPRC of 0.697 (95% CI, 0.653–0.738) and macro F1-score of 0.626 (95% CI, 0.574–0.659). Macro specificity was high at 0.990 (95% CI, 0.989–0.991), while macro sensitivity was 0.573 (95% CI, 0.526–0.618). ST elevation remained challenging, with an F1-score of 0.333 despite an AUROC of 0.854, indicating that ranking performance exceeded fixed-threshold classification in this enriched temporal set (Supplementary Tables 12 and 13).

A complementary public-image evaluation on 103 heterogeneous ECG images from *Life in the Fast Lane* (https://litfl.com) assessed model performance across a substantially different image domain. The model achieved a macro AUROC of 0.962 (95% CI, 0.936–0.983), macro AUPRC of 0.832 (95% CI, 0.770–0.917) and macro F1-score of 0.658 (95% CI, 0.582–0.712). ST elevation could not be estimated because no positive ST elevation cases were present in the selected LITFL set. Compared with the internal test set, AUROC was broadly preserved. VT/VF had a lower point estimate in LITFL but did not remain statistically significant after Holm adjustment, whereas LBBB showed higher AUROC in LITFL after Holm adjustment (Supplementary Tables 14 and 15).

## 3 Discussion

We developed and evaluated an image-based DL method for multi-label ECG classification in emergency care. InCor-EMG combined 18,519 real emergency ECG images, annotations from 19 cardiologists, independent dual review, adjudication, a 12-category emergency ECG taxonomy and comparison with both annotating cardiologists and Mortara Veritas commercial ECG software. The final ConvNeXt ensemble achieved held-out performance close to the mean performance of annotating cardiologists under the adjudicated reference standard and higher F1-scores than mapped Mortara Veritas outputs across most evaluated categories. The model also retained informative performance across scanned and photographed ECGs, with supportive performance in model-enriched temporal and heterogeneous public-image evaluations. These findings support ECG-image classification as a practical route for automated ECG analysis when digital waveforms are unavailable.

The main contribution of this study is the clinically grounded evaluation of ECG-image AI in an emergency-care setting. Most automated ECG systems are designed for digitally stored waveforms, whereas many clinically relevant ECGs circulate as printed tracings, scanned reports, PDFs or mobile photographs. Image-based classification therefore addresses a different deployment problem. It is not intended to replace waveform-based analysis when raw signals are available, but to enable automated analysis when ECG information exists only in visual form. This distinction matters because the visual ECG domain includes sources of variability that are not present in waveform datasets, including layout differences, grid appearance, resolution, scanning artifacts, photography artifacts and cropped or reformatted reports.

The expert-adjudicated structure of InCor-EMG was central to the study. Emergency ECG reading is inherently heterogeneous, and the observed variation in inter-reader agreement was clinically informative. Some ECG categories, such as pacemaker rhythm, premature beats and atrial fibrillation, showed substantial to almost perfect agreement and strong model performance. Others, particularly broad morphologic categories such as ST elevation and the residual Other category, showed lower agreement and greater model difficulty. This distinction was informative for ST-segment abnormalities: in this study, ST elevation captured a broad J-point or ST-segment elevation pattern, whereas injury current was reserved for ECGs expected to trigger immediate reperfusion management. The narrower, emergency-actionable injury current category showed higher agreement and stronger model performance than broad ST elevation, suggesting that clinically aligned target definitions can improve label reproducibility. Across categories, model performance was more closely associated with inter-reader agreement than with training sample size, although inter-reader agreement should be interpreted as a proxy for the underlying visual and labeling ambiguity of each ECG category rather than as an independent causal determinant. The same pattern was observed within ECG categories: AUROC was consistently higher among ECGs for which the two primary cardiologist readers agreed and lower among ECGs that required adjudication. These findings suggest that model errors concentrate near visual and labeling boundaries that are also difficult for human readers. They also indicate that, for poorly reproducible ECG categories, increasing dataset size alone may be insufficient unless the target definition and reference standard are also strengthened. In this setting, dataset quality, adjudication and label reproducibility may be as important as scale.

The comparisons with cardiologists and Mortara Veritas provide complementary benchmarks for the proposed approach. The cardiologist comparison placed model performance in the context of the same expert annotation process used to define the reference standard. Under this adjudicated benchmark, the model achieved macro F1-score close to the mean cardiologist performance, with stronger point estimates for selected rhythm, conduction and emergency-actionable categories, and weaker performance for categories requiring more nuanced visual assessment. The Mortara comparison addressed a different question: whether an image-native model could capture emergency-relevant ECG categories beyond mapped commercial software statements. The model achieved higher F1-scores than mapped Mortara outputs across most evaluated categories, including several acute targets. Together, these comparisons suggest that ECG-image models can provide clinically meaningful structured ECG-category outputs when evaluated against an expert-adjudicated emergency taxonomy.

The additional image-shift analyses further support the relevance of image-based ECG classification for workflows in which ECGs are available only as visual documents. Performance decreased modestly when ECGs were scanned or photographed, but ranking performance remained high and no class-wise F1-score or AUROC decrease remained statistically significant after Holm adjustment. In the model-enriched temporal emergency evaluation, the model retained high AUROC and high specificity despite lower fixed-threshold sensitivity and macro F1-score. However, this result should be interpreted as evidence of informative ranking within an enriched future GE-MUSE image-domain evaluation, not as proof of performance in an unselected prospective emergency workflow. The observed temporal-cohort metrics may reflect case enrichment, reference-standard differences and temporal, device and layout shift. The heterogeneous Life in the Fast Lane evaluation provided an additional public-image stress test, but its small size, curated educational origin and absence of ST elevation positives limit conclusions about generalizability to acute ischemic patterns. Overall, these results suggest that the model was not restricted to the original digital export format, while also emphasizing that operating thresholds should be selected for the intended use case.

These findings have practical implications for emergency ECG AI. ECG-image classification is best viewed as an access-enabling strategy for situations in which the ECG is available visually but not as a digital waveform. In this setting, the relevant question is not whether image-based models should replace waveform-based models, but whether they can recover clinically useful structured information from ECGs that would otherwise be inaccessible to signal-based algorithms. The present results suggest that this is feasible for several emergency-relevant ECG categories, especially when the categories are visually reproducible and clinically well defined.

This study has limitations. The primary dataset came from a single tertiary cardiovascular center, which may limit generalizability across institutions, devices, ECG layouts, patient populations and emergency-care workflows. Although the study included acquisition-shift, model-enriched temporal and public-image evaluations, it did not include a large independent external clinical cohort with the same expert-adjudicated taxonomy. The temporal emergency evaluation was enriched through preselection by an earlier image-based model and labeled by a single cardiologist, and should not be interpreted as an unselected prospective deployment cohort. This design may introduce spectrum bias because the cohort was enriched for ECGs classified as Other and negative for AF and Normal by the earlier model. Therefore, prevalence-sensitive metrics such as AUPRC, F1-score, sensitivity and PPV should be interpreted within this enriched case spectrum. The lower performance observed in this temporal cohort cannot be attributed solely to temporal, device or layout shift because the reference standard also differed from the expert-adjudicated InCor-EMG internal test set. The cardiologist comparison used annotating cardiologists and an adjudicated standard derived from the same annotation process, which provides an annotator-consistency benchmark but does not replace an independent reader study. The Mortara comparison depended on mapping commercial textual statements to the study categories and should be interpreted as a pragmatic comparison with mapped commercial outputs rather than as a direct comparison between systems designed for the same label space. The LITFL evaluation used a small, curated educational image set and lacked positive examples for ST elevation, limiting conclusions about external performance for acute ischemic patterns. Some categories had limited positive sample sizes or greater label uncertainty, particularly VT/VF and broad ST-segment categories, resulting in wider uncertainty or lower fixed-threshold performance.

In conclusion, image-based DL can classify emergency ECGs directly from visual representations with performance close to annotating cardiologists under an adjudicated reference standard and higher F1-scores than mapped commercial ECG outputs across most evaluated categories. The approach remained informative under common visual acquisition shifts and showed supportive performance in model-enriched temporal and heterogeneous public-image evaluations. More broadly, the results indicate that visual and label ambiguity, as reflected by inter-reader agreement, is an important determinant of model performance in emergency ECG-image AI. These findings support ECG-image classification as a promising access-enabling strategy for automated ECG analysis when digital waveforms are unavailable, while emphasizing the need for broader external validation, operating-point selection and prospective workflow evaluation before deployment.

## 4 Methods

### 4.1 Study design and ethics

We conducted a retrospective study of ECGs acquired in the emergency department of a tertiary cardiovascular center between 2018 and 2022. The study was approved by the institutional research ethics committee (CAAE 45070821.3.0000.0068), with waiver of informed consent due to its retrospective design and use of de-identified data.

### 4.2 InCor-EMG cohort assembly

Emergency ECG examinations acquired between 2018 and 2022 were retrieved from the institutional archive of a tertiary cardiovascular center. ECGs were exported in DICOM format from Mortara ELI 250c devices and analyzed as images to match the input format used by the proposed model. A representative full ECG image in this format is shown in Supplementary Figure 1. The cohort was restricted to adults aged 18 years or older. ECGs labeled as *Exclusion* during expert review, including technically invalid, severely artifacted, lead-inverted or unreadable tracings, were removed from the analytical cohort. After filtering, 18,519 ECGs from 14,402 unique patients were retained for analysis. ECGs were represented as three-channel images with dimensions of 3122 by 1671 pixels.

### 4.3 Emergency ECG taxonomy and reference standard

A panel of cardiologists identified 19 candidate ECG findings relevant to emergency ECG analysis and an additional category for technically invalid or unclassifiable exams. These findings were consolidated into ECG categories according to clinical similarity and relevance to acute decision making (Supplementary Table 1). The final modeled target space included 12 acute ECG categories: Normal, Atrial Fibrillation (AF), Atrial Tachycardia or Atrial Flutter (AT/Flutter), Paroxysmal Supraventricular Tachycardia or Sinus Tachycardia (PSVT/ST), Right Bundle Branch Block (RBBB), Left Bundle Branch Block (LBBB), Second-degree, Third-degree or Advanced Atrioventricular Block or Sinus Bradycardia (AVB 2°/3°/advanced/SB), Premature beat, Pacemaker rhythm (PM), ST elevation, Injury current and Ventricular Tachycardia or Ventricular Fibrillation (VT/VF). Representative ECG patterns from leads II, V1 and V5 for each modeled target class are shown in Supplementary Figure 2.

ECGs were treated as multi-label instances. *Normal* and *Exclusion* were mutually exclusive labels. ECGs without any modeled target-class finding were assigned to *Other*. This category was not intended to represent a homogeneous ECG finding, but rather a residual group of non-target abnormalities and ECGs outside the selected emergency taxonomy. The *Exclusion* label captured technical invalidity and was removed from the analytical cohort. All remaining target classes followed a multi-label structure.

#### 4.3.1 Class-specific annotation criteria

Class-specific annotation criteria were based on the Brazilian Society of Cardiology guideline for ECG analysis and reporting whenever applicable [13]. The cardiologist panel defined additional operational rules for grouped classes, visually ambiguous findings and emergency-actionable patterns before labeling. The annotation guidelines were shared with all participating physicians. The operational criteria were as follows:

##### Normal

Normal ECG was annotated when no modeled target-class finding was present and the tracing was considered within normal limits for the emergency ECG taxonomy. Normal was mutually exclusive and could not coexist with any other ECG category.

##### Atrial fibrillation (AF)

AF required an irregularly irregular RR pattern and fibrillatory activity. AF could coexist with right or left bundle branch block, premature beats and pacemaker capture.

##### Atrial tachycardia or atrial flutter (AT/Flutter)

AT/Flutter included organized atrial tachycardias and typical or atypical atrial flutter patterns.

##### Paroxysmal supraventricular tachycardia or sinus tachycardia (PSVT/ST)

PSVT/ST was defined as an operational ECG category including paroxysmal supraventricular tachycardia, atrioventricular nodal reentrant tachycardia, atrioventricular reentrant tachycardia and sinus tachycardia. Sinus tachycardia was annotated only when the rhythm was unequivocally sinus and at least 17 QRS complexes were present in 10 seconds in the long lead II rhythm strip.

##### Right bundle branch block (RBBB)

RBBB followed established ECG criteria for right bundle branch block.

##### Left bundle branch block (LBBB)

LBBB followed established ECG criteria for left bundle branch block. In the presence of LBBB, suspected left ventricular hypertrophy was labeled as *Other* because of label uncertainty.

##### Second-degree, third-degree or advanced atrioventricular block or sinus bradycardia (AVB 2°/3°/advanced/SB)

AVB 2°/3°/advanced/SB was defined as an operational emergency ECG category including Mobitz I, Mobitz II, 2:1 atrioventricular block, high-grade atrioventricular block, complete atrioventricular block and extreme sinus bradycardia. Extreme sinus bradycardia was annotated when 7 or fewer QRS complexes were present in 10 seconds. First-degree atrioventricular block was not included in this ECG category and was labeled as *Other*. Paced rhythms with ventricular capture were labeled only as *Pacemaker rhythm*, even if atrioventricular dissociation was present.

##### Premature beat

Premature beat included atrial and ventricular ectopy occurring as isolated beats, pairs or triplets, and also non-sustained ventricular tachycardia.

##### Pacemaker rhythm (PM)

PM was annotated whenever pacing spikes with ventricular capture were visible. PM could coexist with AF, atrial flutter and premature beats.

##### ST elevation

ST elevation followed the general criterion of J-point or ST-segment elevation in at least two contiguous leads, with operational thresholds of *≥*1 mm in limb and left precordial leads and *≥*2 mm in V1–V3. Thresholds were not adjusted for age or sex. In ventricular pre-excitation, ST elevation was not assigned and pre-excitation was recorded as *Other*. In paced rhythms, ST elevation was not assigned.

##### Injury current

Injury current was reserved for ECGs expected to trigger immediate reperfusion management. In LBBB, a Sgarbossa score *≥*5 served as supportive evidence.

##### Ventricular tachycardia or ventricular fibrillation (VT/VF)

A conservative rule was applied for VT/VF. Any regular wide-QRS tachycardia with QRS duration *≥*120 ms was labeled as ventricular tachycardia. Ventricular fibrillation was reserved for chaotic tracings without discernible QRS complexes.

### 4.4 Annotation workflow and inter-reader agreement

InCor-EMG was divided into 20 annotation batches of approximately 1,000 ECGs. Each ECG was independently reviewed by two cardiologists using a secure web-based annotation platform. The platform displayed the ECG image, patient age and sex, and the complete list of ECG classes (Supplementary Figure 3). Each annotator accessed the platform through an individual login, enabling traceability of labels, review actions and adjudication decisions.

Disagreements in modeled target ECG categories were adjudicated by a third cardiologist to define the final reference standard. Nineteen cardiologists participated in total. The full annotation and adjudication workflow is shown in Supplementary Figure 4.

Inter-reader agreement between the two primary annotators was quantified for each ECG class using Cohen’s *κ*. Nonparametric 95% confidence intervals were computed using ECG-level bootstrap resampling with 1,000 iterations, preserving the multi-label structure within each resampled ECG. Agreement categories followed conventional agreement thresholds [14].

### 4.5 ECGIMGSin and InCor-AMB dataset construction

Two additional ECG image datasets were used before final model development on InCor-EMG. ECGIMGSin was a synthetic ECG image dataset generated from public ECG waveform datasets, including CODE15 and CINC21 [15, 16]. Because these datasets are distributed primarily as time-series signals, waveforms were rendered as paper-like ECG images using an adapted ECG image generation pipeline [17]. Labels were harmonized to the six CODE15-compatible classes: first-degree atrioventricular block, right bundle branch block, left bundle branch block, sinus bradycardia, atrial fibrillation and sinus tachycardia.

Clinical ECG printouts may vary in lead arrangement, number of columns and presence of long rhythm strips. We therefore generated synthetic images using multiple layouts, including four-column, two-column and single-column formats, with different long-lead configurations. This was intended to reduce dependence on a single visual organization and expose the model to ECG image formats commonly encountered in clinical practice. To increase image-domain variability, the generation pipeline introduced visual augmentations including grid variation, image noise, rotation, paper-like wrinkles, random resolution and optional header insertion. These augmentations were intended to approximate visual differences encountered in exported, scanned or photographed ECG images. Supplementary Figure 6 illustrates the synthetic ECG-image generation pipeline.

InCor-AMB was a retrospective ambulatory ECG image dataset from the *Instituto do Coração do Hospital das Clínicas da Faculdade de Medicina da Universidade de São Paulo* (InCor-HCFMUSP), previously described by Dias et al. [12]. ECGs acquired between 2017 and 2020 were retrieved from institutional DICOM archives with available demographic metadata and clinical reports. Exams from patients younger than 18 years and exams without an associated report were excluded. All exams were acquired on Mortara ELI 250c devices. ECG images were extracted from DICOM files, converted to image format and cropped to remove sensitive patient information while preserving the ECG tracing. For consistency with ECGIMGSin, only the six CODE15-compatible categories were used at this stage.

These datasets were used for pretraining and intermediate model development, whereas the final emergency target space and final evaluation were defined using InCor-EMG.

### 4.6 Data splitting

A patient-level split reserved 20% of InCor-EMG as a held-out internal test set, while the remaining 80% was used for model development through 10-fold cross-validation. Patient-level separation was enforced across all splits to avoid information leakage from repeated ECGs from the same patient. The full data-splitting scheme, including the cross-validation structure used within the development set, is illustrated in Supplementary Figure 5.

### 4.7 Architecture selection and training optimization

Architecture selection and training optimization were performed using only the InCor-EMG development set. All architecture-selection and optimization experiments used 10-fold cross-validation within the development set, with patient-level separation preserved across folds. The held-out test set was not used for architecture selection, optimization, threshold selection or any other model-development decision. This development framework extended our preliminary ECG-image classification work with pre-trained ConvNeXt models from the 2024 PhysioNet/CinC Challenge [18].

#### 4.7.1 Candidate architectures

We evaluated five candidate image-classification backbones: ConvNeXt, EfficientNet-B3, ResNet50, Vision Transformer (ViT) and Swin Transformer (SwinT) [19–23]. These architectures were selected to represent different families of modern computer vision models, including convolutional neural networks, hybrid hierarchical models and transformer-based architectures. For the initial architecture-selection stage, all backbones were trained using the same baseline training protocol and evaluated on the 10 validation folds of the InCor-EMG development set. Performance was summarized using the mean class-wise F1-score across the 12 modeled ECG classes. The architecture with the highest mean F1-score was selected for all subsequent optimization experiments.

#### 4.7.2 Baseline training configuration

The baseline configuration used ImageNet-initialized backbone weights [24, 25] and fine-tuned each candidate architecture on the InCor-EMG multi-label classification task. ECG images were resized to 224 x 224 pixels. Although the final emergency taxonomy comprised 12 modeled ECG classes, the training head included 13 independent outputs: the 12 target classes and one auxiliary output defined as the union of ST elevation and injury current. This auxiliary label was positive when either ST elevation or injury current was present, was intended to support learning of shared ischemic ECG features, and was excluded from all reported class-wise, macro-averaged, cardiologist-comparison and Mortara-comparison performance analyses.

The image preprocessing pipeline converted ECG images to grayscale, applied contrast-limited adaptive histogram equalization and normalized pixel intensities using ImageNet mean and standard deviation before tensor conversion. During training, image-domain augmentations were applied to improve robustness to visual variability in ECG images. These included random brightness and contrast adjustment, Gaussian noise, shift-scale-rotation, elastic transformation and grid distortion. The same deterministic preprocessing, without random augmentations, was used for validation. Models were trained with binary cross-entropy with logits using the AdamW optimizer [26]. The learning rate was fixed at 1 *×* 10*^−^*^4^, with no learning-rate scheduler. Training used a batch size of 64, mixed precision, gradient clipping with maximum norm 1.0 and a maximum of 30 epochs. Early stopping monitored validation F1-score with patience of 6 epochs and a minimum improvement of 0.001. The checkpoint with the best validation F1-score was restored for each fold.

#### 4.7.3 Data augmentation

We evaluated image-domain data augmentation as a strategy to improve robustness to visual variability in ECG images. Augmentations were applied only during training and were designed to preserve ECG morphology and category-relevant content while increasing variability in image appearance. These transformations included brightness and contrast variation, Gaussian noise, small translations, small rotations, elastic deformation and grid distortion. Transformations that could alter the physiological or temporal structure of the ECG tracing, such as horizontal flipping, were not used.

#### 4.7.4 ECG-domain pretraining

We evaluated ECG-domain pretraining using ECGIMGSin and InCor-AMB before fine-tuning on InCor-EMG. ECGIMGSin provided a large synthetic ECG image corpus generated from public ECG waveform datasets, whereas InCor-AMB provided real ambulatory ECG images from the same institution. This pretraining stage was intended to expose the network to ECG-specific visual structures, including waveform morphology, grid patterns, lead layouts and image-domain artifacts, before training on the emergency ECG target task.

Two ECG-domain pretraining variants were assessed. In the first variant, the network was initialized with random weights and pretrained on ECGIMGSin and InCor-AMB before fine-tuning on InCor-EMG. In the second variant, the network was initialized with ImageNet weights, further pretrained on ECGIMGSin and InCor-AMB, and then fine-tuned on InCor-EMG. During fine-tuning, all layers were left trainable rather than freezing early layers.

#### 4.7.5 Metadata integration

We evaluated whether patient metadata improved ECG image classification. Age and sex were incorporated as additional non-image inputs. Sex was encoded as a binary variable, with male encoded as 1 and female or other/unknown encoded as 0. Age was normalized by dividing the value in years by 100. These metadata inputs were concatenated with the image-derived feature vector before the final classification layer.

#### 4.7.6 Custom loss function

Because InCor-EMG is a multi-label dataset with substantial class imbalance, we evaluated a custom loss function designed to complement binary cross-entropy with a metric-aware term. The custom loss combined binary cross-entropy with a differentiable approximation of the Matthews correlation coefficient (MCC), which accounts for true positives, true negatives, false positives and false negatives.

The standard MCC is defined as:

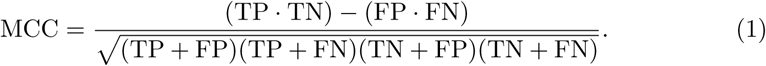

To make MCC differentiable, binary counts were replaced by soft probabilistic approximations computed from predicted probabilities and reference labels. The custom loss was defined as:

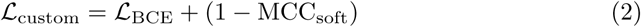

#### 4.7.7 High-resolution retraining

We evaluated high-resolution retraining to determine whether finer ECG morphology improved classification performance. In this strategy, the model was first trained using ECG images resized to 224 x 224 pixels. The resulting weights were then used to initialize a second training stage using the same ECG images resized to 384 x 384 pixels. This two-stage procedure was intended to combine the stability and computational efficiency of lower-resolution training with the additional morphological detail available at higher resolution.

#### 4.7.8 Ablation design

The optimization strategies were evaluated through a structured ablation study using the selected backbone. Ten experimental configurations were assessed:

- **Experiment 0:** Baseline model initialized with ImageNet weights, without additional optimization.
- **Experiment 1:** Baseline model with data augmentation.
- **Experiment 2:** ECG-domain pretraining on ECGIMGSin and InCor-AMB initialized from random weights, followed by fine-tuning on InCor-EMG.
- **Experiment 3:** ImageNet initialization followed by ECG-domain pretraining on ECGIMGSin and InCor-AMB, followed by fine-tuning on InCor-EMG.
- **Experiment 4:** Baseline model with metadata integration using age and sex.
- **Experiment 5:** Baseline model trained with the custom loss function.
- **Experiment 6:** Baseline model with high-resolution retraining from 224 x 224 pixels to 384 x 384 pixels.
- **Experiment 7:** Combination of data augmentation and ImageNet plus ECG-domain pretraining.
- **Experiment 8:** Combination of data augmentation, ImageNet plus ECG-domain pretraining and the custom loss function.
- **Experiment 9:** Combination of data augmentation, ImageNet plus ECG-domain pretraining, the custom loss function and high-resolution retraining.

For each experiment, performance was computed on each of the 10 validation folds and summarized as the mean class-wise F1-score across the 12 modeled ECG classes. The final optimized configuration was selected as the experiment with the highest mean F1-score across the 10 folds. The selected configuration was then used to train the final ensemble, consisting of the 10 cross-validation models, with final predictions obtained by averaging predicted probabilities across ensemble members.

### 4.8 Internal test evaluation and comparator analyses

The final model was evaluated on the held-out InCor-EMG test set, which was not used during architecture selection, training optimization, threshold selection or any other model-development step. The final model consisted of an ensemble of the 10 optimized ConvNeXt models trained during 10-fold cross-validation on the InCor-EMG development set. For each ECG, the predicted probability for each ECG class was computed as the average probability across the 10 ensemble members.

The final model was compared with the cardiologists who annotated InCor-EMG using the adjudicated label as the reference standard. Cardiologist performance was computed by comparing each available independent reader annotation with the final adjudicated label for the same ECG. When both independent reader annotations were available for an ECG, both were included while accounting for clustering by ECG during uncertainty estimation.

Commercial ECG classification was evaluated using Mortara Veritas^™^ reports available for a subset of the held-out InCor-EMG test set. A cardiologist mapped the textual Mortara Veritas^™^ statements to the modeled ECG categories before analysis. These mapped labels were compared with the adjudicated InCor-EMG reference standard and with the predictions of the final model on the same ECGs.

### 4.9 Exploratory class-level analysis of performance determinants

We performed exploratory class-level analyses to assess whether between-class variation in model performance was more closely associated with annotation consistency or training sample size. Each ECG category was treated as one analytical unit. Inter-reader agreement was quantified using the class-wise Cohen’s *κ* between the two primary cardiologist readers, and training sample size was quantified as the log10-transformed number of positive training examples in the development set. Class-level model performance was evaluated using AUROC and prevalence-normalized AUPRC. Normalized AUPRC was computed as (AUPRC *− π*)*/*(1 *− π*), where *π* denotes the class prevalence in the test set.

### 4.10 Performance in scanned and photographed ECG images

To evaluate performance in acquisition shifts commonly encountered in clinical workflows, we constructed a 120-ECG subset from the held-out InCor-EMG internal test set by randomly selecting 10 examples for each modeled ECG category. The same ECGs were evaluated under three image-acquisition conditions: original digital ECG images, digitally scanned printed ECGs and smartphone photographs of printed ECGs. The adjudicated InCor-EMG labels were used as the reference standard for all three conditions, ensuring that comparisons across acquisition formats were paired at the ECG level.

The final 10-fold ConvNeXt ensemble was applied to each image version without retraining or threshold recalibration. To quantify degradation relative to the original digital format, paired class-wise differences were computed between original digital images and each acquisition-shift condition. Positive deltas indicate higher performance on original digital images.

### 4.11 Enriched temporal emergency evaluation

We performed a model-enriched temporal emergency evaluation using future ECGs acquired after the InCor-EMG study period. The source dataset consisted of emergency ECG images acquired in 2023–2024 from a GE-MUSE system, whereas the InCor-EMG training and internal test ECGs were exported from Mortara ELI 250c devices. This evaluation therefore introduced a combined temporal, device and layout shift.

This cohort was not intended to represent an unselected prospective emergency validation cohort. Future ECGs were first processed by the previously described Dias et al. model [12], which classifies ECG images as AF, Normal or Other. ECGs classified as Other and negative for both AF and Normal by that earlier model were selected for the present evaluation. The proposed model and its predictions were not used for case selection. This design enriched the temporal cohort for non-normal, non-AF ECGs requiring finer emergency-category classification.

The selected ECGs were labeled by a single cardiologist in 2025 using the same ECG category criteria applied in InCor-EMG. ECGs labeled as *Exclusion* were removed from performance analyses. After exclusion of technically invalid or non-evaluable ECGs, 1,855 images were included in the evaluation. Predictions were generated using the final 10-fold ConvNeXt ensemble without retraining, threshold recalibration or any additional adaptation to the temporal data.

Because the cohort was enriched through a previous image-based model and labeled by a single cardiologist rather than independently reviewed and adjudicated, this analysis was treated as a model-enriched temporal stress test rather than as an unselected or expert-adjudicated validation cohort. To contextualize performance in this enriched future image domain, class-wise AUROC in the model-enriched temporal evaluation was compared with AUROC in the held-out InCor-EMG internal test set.

### 4.12 External evaluation on heterogeneous web ECG images

External evaluation was performed using ECG images from the Life in the Fast Lane (LITFL) repository. This dataset was used to assess model generalization to heterogeneous public ECG images that differ from the institutional data in layout, resolution, graphical style and acquisition source. The LITFL set included 103 ECG images, each treated as an independent image-level unit.

Reference labels were harmonized to the same multi-label ECG taxonomy used for InCor-EMG. The final 10-fold ConvNeXt ensemble was applied to the LITFL images without retraining, domain adaptation or threshold recalibration. Classes with no positive examples in LITFL were retained in prevalence summaries but were considered non-estimable for metrics requiring positive cases. Class-wise AUROC in LITFL was compared with AUROC in the held-out InCor-EMG internal test set as an unpaired cross-cohort comparison.

### 4.13 Statistical analysis

Model performance was evaluated under a multi-label formulation using the reference labels defined for each evaluation set. For each ECG category, we computed the area under the receiver operating characteristic curve (AUROC), area under the precision-recall curve (AUPRC), sensitivity (Se), specificity (Spe), positive predictive value (PPV) and F1-score. Predicted probabilities were converted into binary predictions using a fixed threshold of 0.5 for all ECG categories. Macro-averaged metrics were computed as the unweighted mean of class-wise metrics. Class-level values that were not estimable because of absent positive cases or zero predicted positives were excluded from the corresponding macro average.

Uncertainty was estimated using nonparametric bootstrap resampling with 1,000 iterations. Patient-level cluster bootstrap resampling was used for the internal test set and enriched temporal emergency evaluation when patient identifiers were available. ECG-level bootstrap resampling was used for acquisition-shift comparisons, preserving the paired structure across acquisition formats. Image-level bootstrap resampling was used for the LITFL evaluation. For the cardiologist comparison, uncertainty was estimated using ECG-level cluster bootstrap resampling to account for multiple reader annotations from the same ECG. For all bootstrap analyses, the multi-label structure of each resampled unit was preserved.

Pairwise comparisons were performed using paired resampling when predictions were available on the same ECGs, including model-versus-cardiologist comparisons, model-versus-Mortara comparisons and acquisition-shift comparisons. Cross-cohort AUROC comparisons, including LITFL versus the internal test set and enriched temporal emergency evaluation versus the internal test set, were treated as unpaired comparisons because the datasets contained different ECGs. P values were obtained from permutation tests or bootstrap metric draws, as appropriate for each comparison, and were adjusted across ECG categories using the Holm method.

For the exploratory class-level analysis of performance determinants, associations between model performance, inter-reader agreement and training sample size were assessed using Spearman correlation and multivariable linear regression with standardized predictors. Model fit was summarized using in-sample *R*^2^ and leave-one-class-out *R*^2^. Bootstrap sensitivity analyses resampled ECG categories with replacement and computed the difference in Spearman correlations between inter-reader agreement and performance versus training sample size and performance. These analyses were considered exploratory and hypothesis-generating.

## Data Availability

The ECG data used in this study were obtained from the Heart Institute (InCor), Hospital das Cĺınicas, University of São Paulo Medical School (HCFMUSP), and are subject to institutional governance, ethical approval and applicable privacy regulations, including the Brazilian Lei Geral de Proteção de Dados (LGPD). The complete individual-level InCor ECG images, clinical metadata and annotations cannot be made publicly available because they derive from personal health data and may contain sensitive information.

A small set of anonymized sample ECG images derived from the InCor database is available at https://github.com/fmenegui/image-based-ecg-classification-emergency to illustrate the data format and support reproducibility of the inference workflow. These samples do not constitute the full study dataset. The repository also provides prediction outputs and evaluation files generated in this study, including model predictions on the InCor-EMG test set, Mortara commercial classification outputs, LITFL external-evaluation predictions and future InCor-EMG predictions, together with the corresponding released labels or identifiers where permitted by governance and source terms.

Requests for access to de-identified InCor-derived data for non-commercial research or educational purposes will be evaluated on a case-by-case basis. Requests should be submitted to Prof. Jose Eduardo Krieger and include the study protocol, institutional affiliation of the investigators, ethics approval or waiver when applicable, a data protection plan, and a detailed description of the requested variables and intended analyses. All requests will undergo institutional and ethical review, subject to applicable governance procedures. If approved, access will be granted under a formal Data Use Agreement, which prohibits data sharing, commercial use, attempts at re-identification, and unauthorized linkage with external datasets.

The Life in the Fast Lane (LITFL) ECG images used for external evaluation were obtained from the publicly accessible LITFL ECG Library. To avoid redistribution of third-party educational material, the reproducibility materials provide source-page links, derived labels and prediction outputs, but do not include the original LITFL images. Users should retrieve any LITFL images directly from their original source and comply with the applicable LITFL terms, including appropriate attribution to litfl.com, non-commercial use and exclusion of images labelled as belonging to third parties.

## Code availability

Code for inference, model evaluation and reproducibility workflows is available at https://github.com/fmenegui/image-based-ecg-classification-emergency. The repository includes scripts for single-image inference, grouped cross-validation training, data-structure checks, examples of the released data schema, anonymized InCor sample ECG images, and reproducibility materials for the evaluation experiments. These materials include prediction outputs for the InCor-EMG test set, Mortara commercial ECG classification, the LITFL external-evaluation set and the future InCor-EMG evaluation, as well as source-page links, derived labels and instructions to retrieve LITFL images from their original source. The repository does not redistribute the original LITFL images or the complete restricted InCor datasets.

The final ConvNeXt ensemble weights are hosted at https://huggingface.co/fmenegui/ecg-image-convnext-ensemble; access may require approval from the authors because the model was developed from restricted clinical data. The ECG-image pretraining checkpoint is hosted at https://huggingface.co/fmenegui/pretrained_image_ecg_doc. A demonstration application is available at https://huggingface.co/spaces/fmenegui/image-based-emergency-ecg-prediction. Documentation in the repository describes how to reproduce inference on compatible ECG images and how to train models on authorized local datasets.

## Acknowledgements

The authors thank the cardiologists who participated in ECG review and adjudication. This research was supported by Foxconn Brazil and the Zerbini Foundation as part of the research initiative titled “Machine Learning in Cardiovascular Medicine” and FAPESP grant 2019/25153-0.

## Author contributions

F.M.D. contributed to conceptualization, data curation, methodology, software development, formal analysis, validation, visualization and writing of the original draft.

E.R. contributed to data curation, methodology, validation and manuscript review and editing. N.O. contributed to clinical supervision, ECG taxonomy definition, clinical validation and manuscript review and editing. O.A.O.C. contributed to ECG annotation, clinical validation and manuscript review and editing. C.A.P. contributed to clinical supervision, ECG taxonomy definition, clinical validation and manuscript review and editing. J.E.K. contributed to institutional supervision, data governance, funding acquisition and manuscript review and editing. M.A.G. contributed to conceptualization, supervision, methodology, funding acquisition, project administration and manuscript review and editing. All authors reviewed and approved the final manuscript.

## Competing interests

The authors declare no competing interests.

## Supplementary Appendix A Supplementary Figures

**Supplementary Figure 1.**
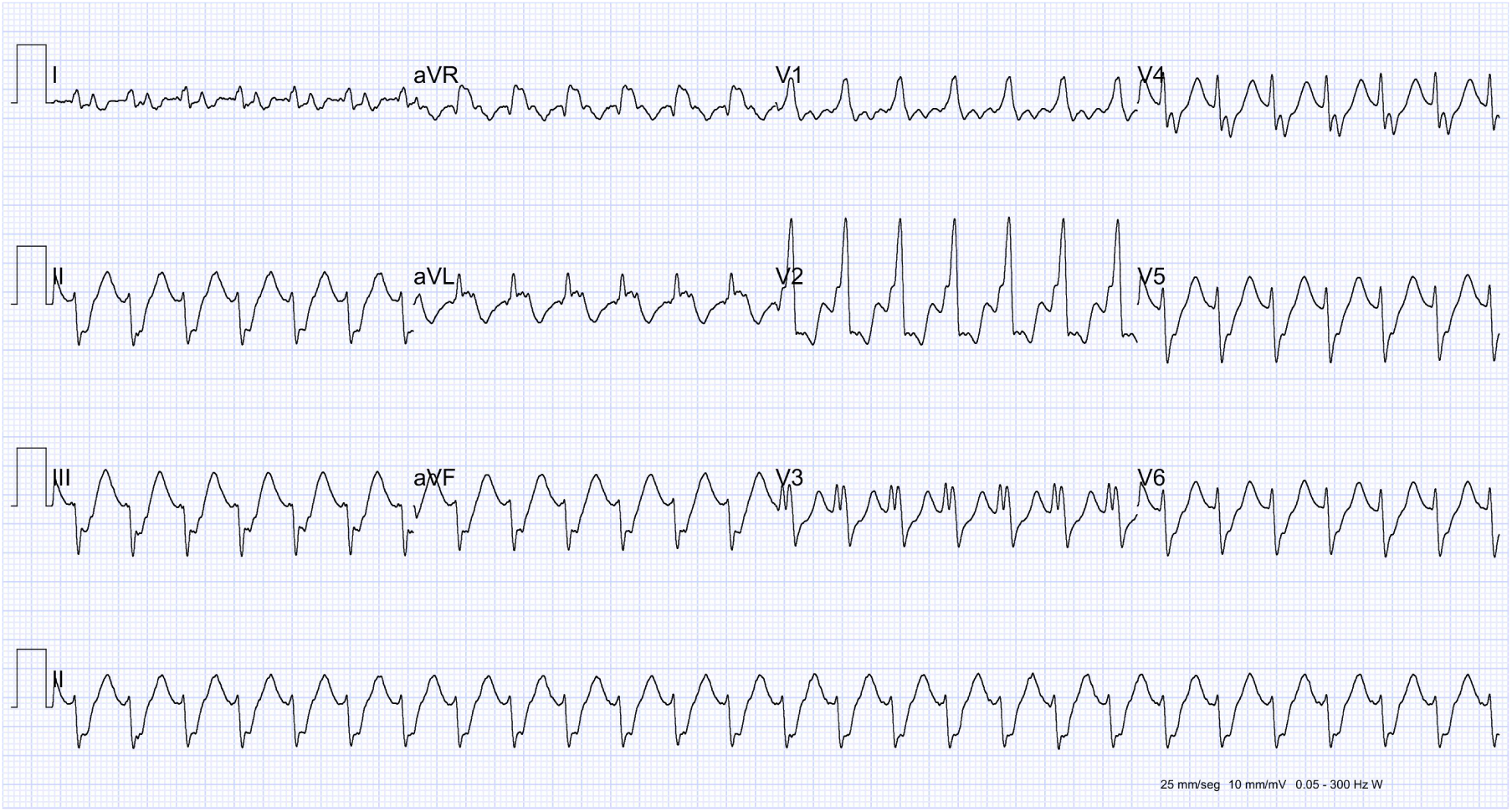
Representative full ECG image from InCor-EMG dataset. ECGs were exported from Mortara ELI 250c devices and represented as three-channel images with dimensions of 3122 by 1671 pixels.

**Supplementary Figure 2.**
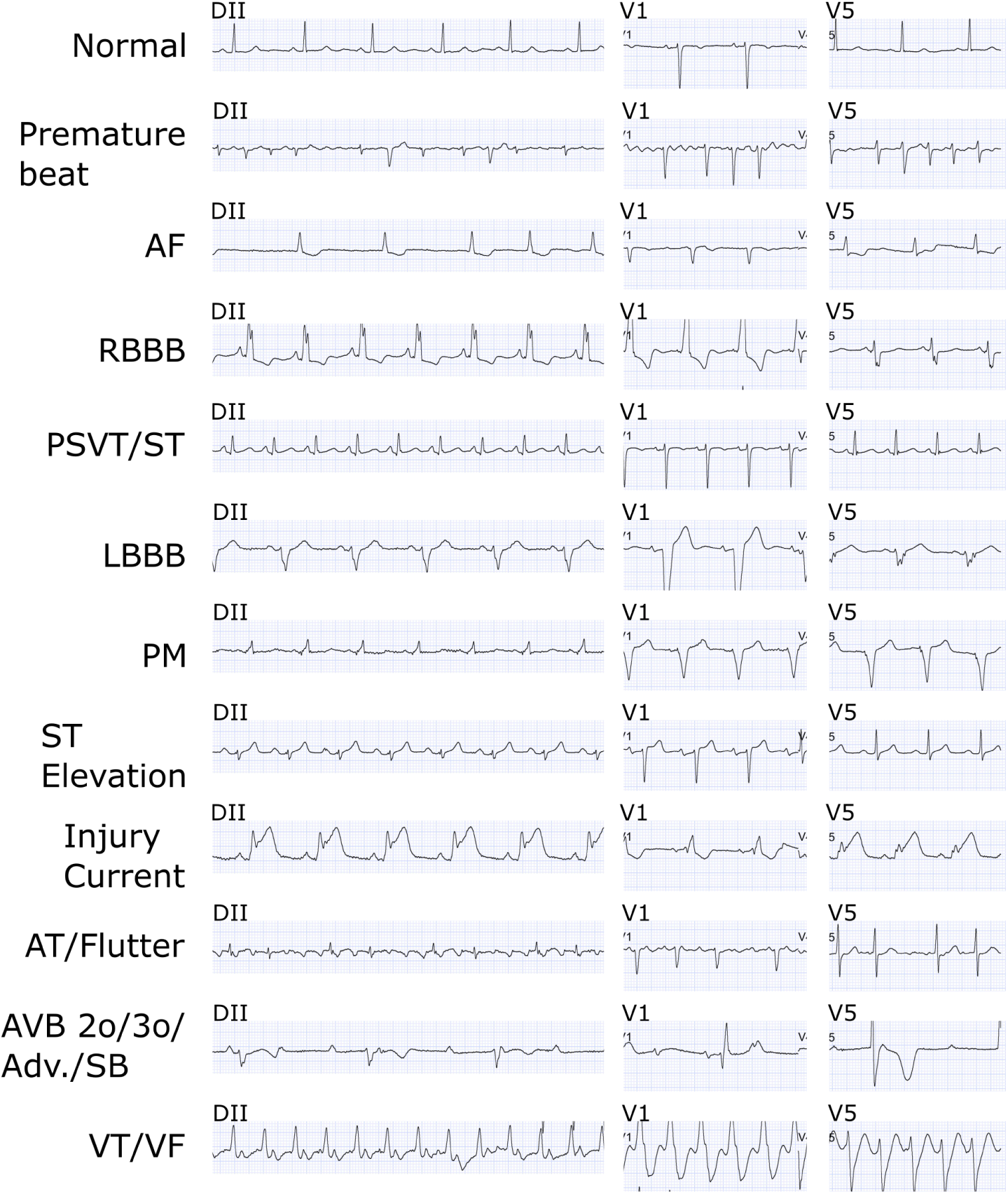
Representative ECG patterns from leads II, V1 and V5 for the 12 modeled InCor-EMG target classes: (a) Normal, (b) Premature beat, (c) Atrial fibrillation, (d) Right bundle branch block, (e) paroxysmal supraventricular tachycardia/sinus tachycardia, (f) left bundle branch block, (g) pacemaker rhythm, (h) ST-segment elevation, (i) injury current, (j) atrial tachycardia/flutter, (k) second-degree, third-degree or advanced atrioventricular block/sinus bradycardia, and (l) ventricular tachycardia/ventricular fibrillation.

**Supplementary Figure 3.**
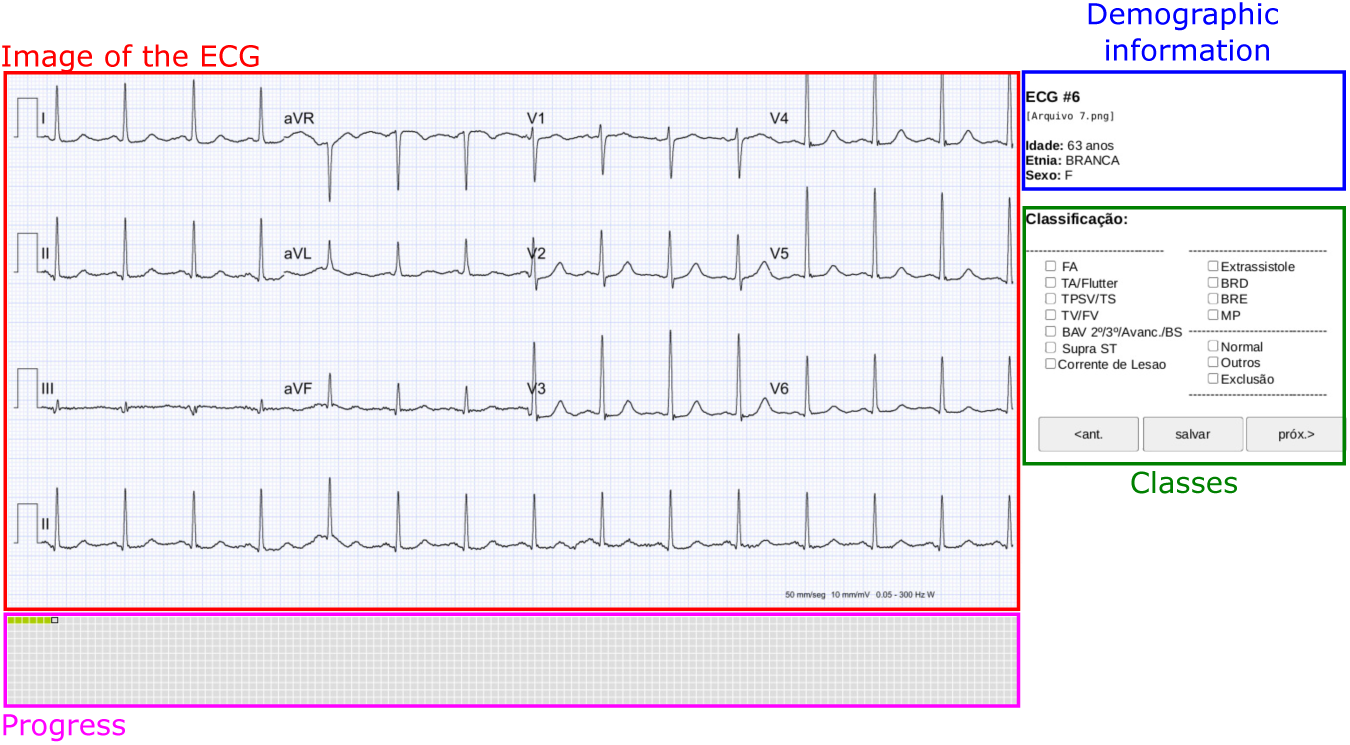
Web-based annotation platform used for InCor-EMG. The platform displayed the ECG image, patient age and sex, and the full list of ECG classes. Individual logins enabled traceability of annotations and adjudication decisions.

**Supplementary Figure 4.**
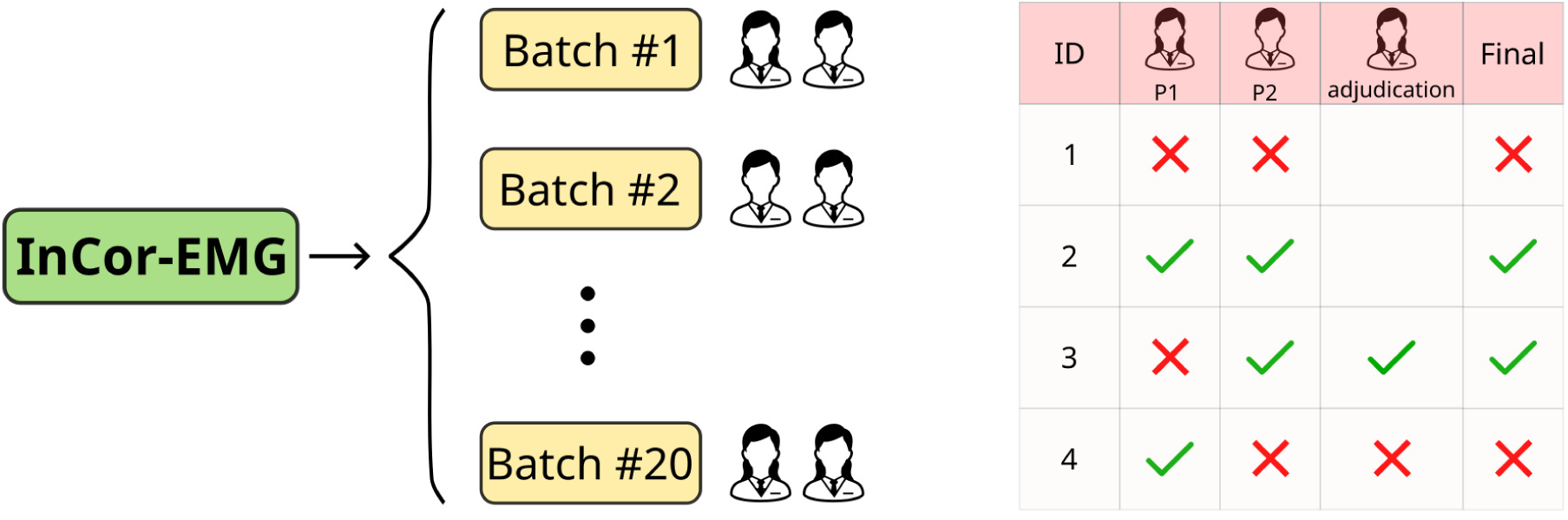
InCor-EMG annotation and adjudication workflow. ECGs were divided into 20 annotation batches. Each ECG was independently reviewed by two cardiologists, and disagreements were adjudicated by a third cardiologist to define the final reference standard.

**Supplementary Figure 5.**
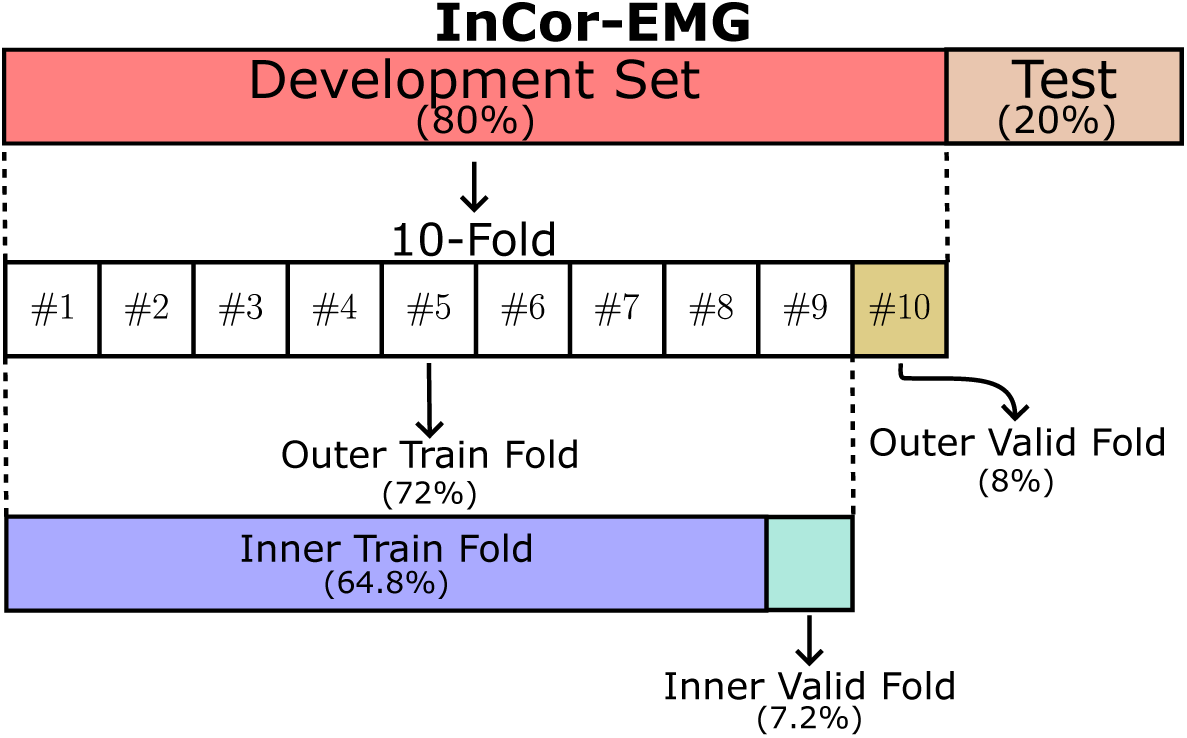
Patient-level data splitting strategy for InCor-EMG. A held-out internal test set included 20% of the cohort, while the remaining 80% formed the development set used for cross-validation and model selection.

**Supplementary Figure 6.**
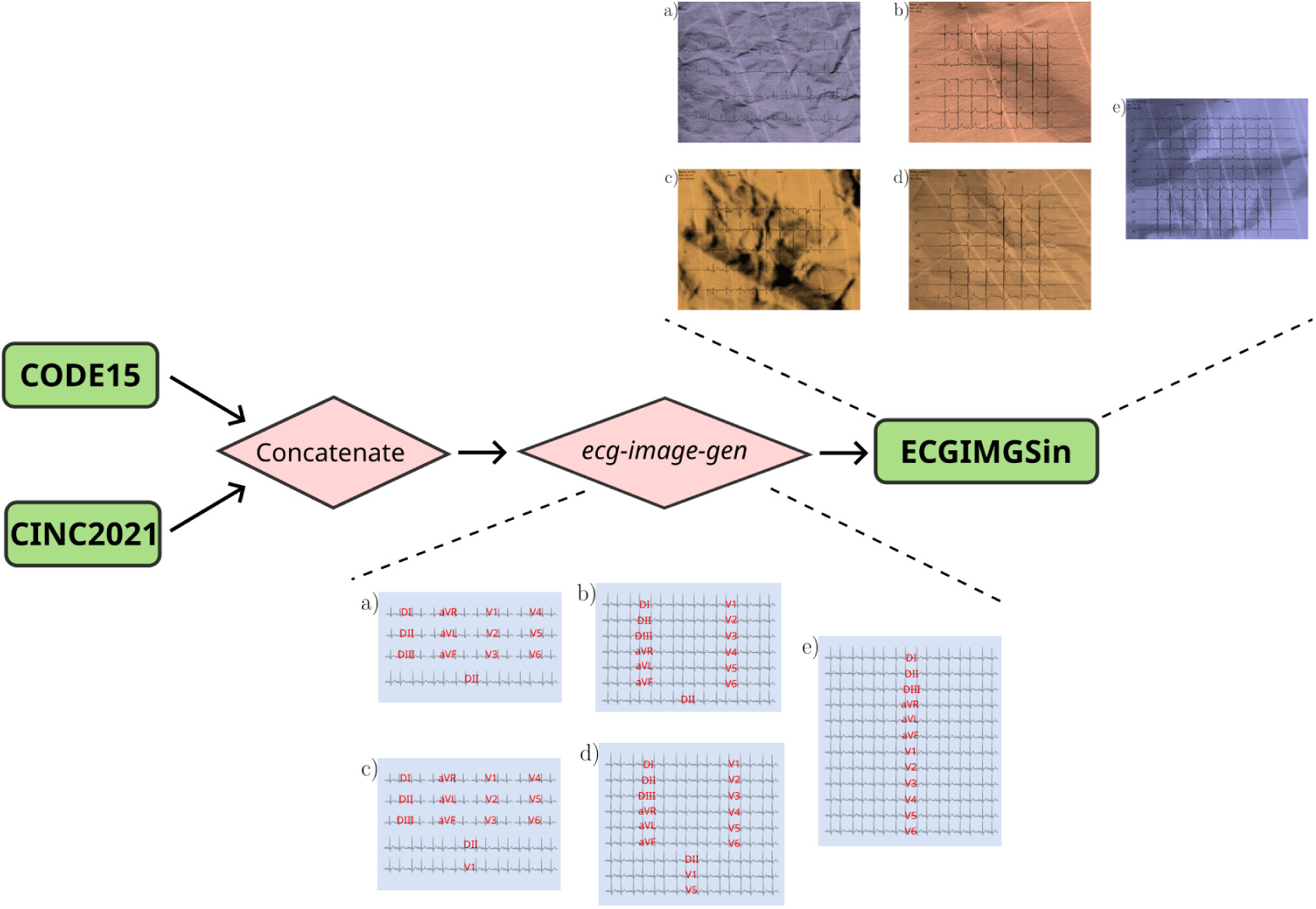
ECGIMGSin synthetic ECG image generation pipeline. Public ECG waveform datasets from CODE15 and CINC21 were harmonized into a shared six-label ECG space and rendered as paper-like ECG images using an adapted ECG image generation pipeline. The generator produced multiple clinical layouts and visual augmentations to increase image-domain variability.

**Supplementary Figure 7.**
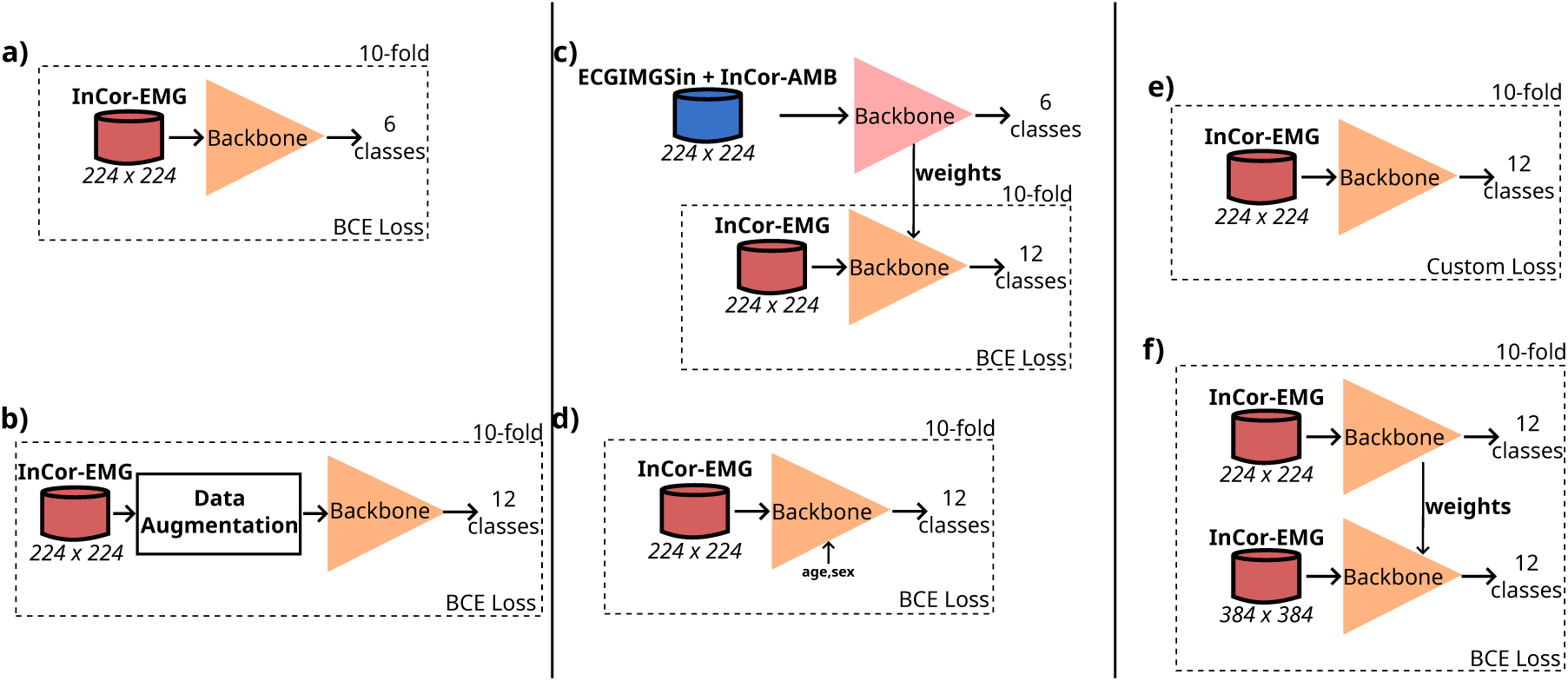
Architecture-selection and optimization workflow used during model development. (a) Baseline architecture-selection experiment, in which candidate backbones were trained on InCor-EMG images resized to 224×224 pixels using binary cross-entropy loss and evaluated by 10-fold cross-validation. (b) Data-augmentation experiment, adding image-domain transformations during training to improve robustness to visual variability. (c) ECG-domain pretraining experiment, in which the backbone was first trained on ECGIMGSin and InCor-AMB and the resulting weights were transferred to the 12-class InCor-EMG task. (d) Metadata-integration experiment, adding age and sex to the image-derived representation before classification. (e) Custom-loss experiment, replacing the baseline binary cross-entropy objective with the custom loss used for optimization. (f) High-resolution retraining experiment, in which weights learned at 224×224 pixels were transferred to a second training stage using 384×384-pixel ECG images. All optimization experiments were performed within the InCor-EMG development set using 10-fold cross-validation.

**Supplementary Figure 8.**
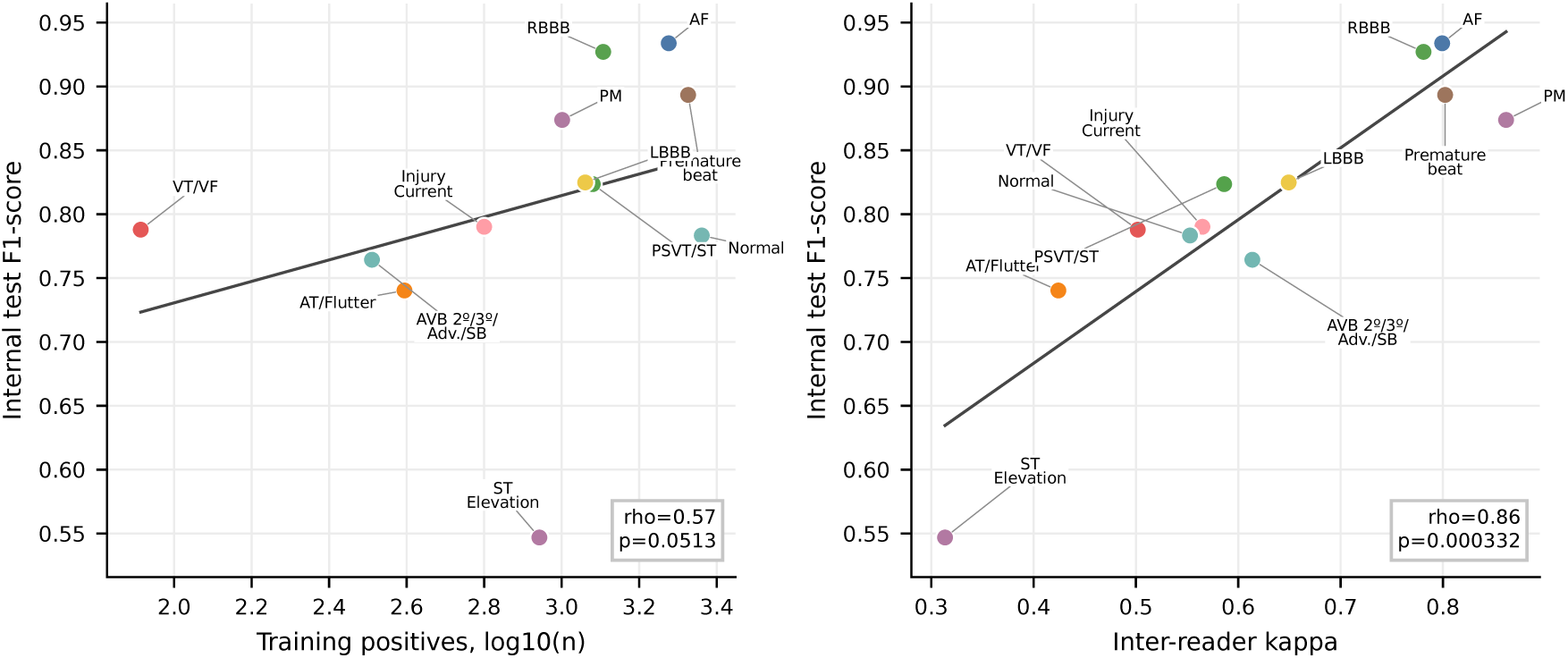
Association between class-wise F1-score, inter-reader agreement and positive training examples. Each point represents one modeled ECG category. Inter-reader agreement was measured using Cohen’s *κ*, and training sample size was represented by the number of positive training examples.

**Supplementary Figure 9.**
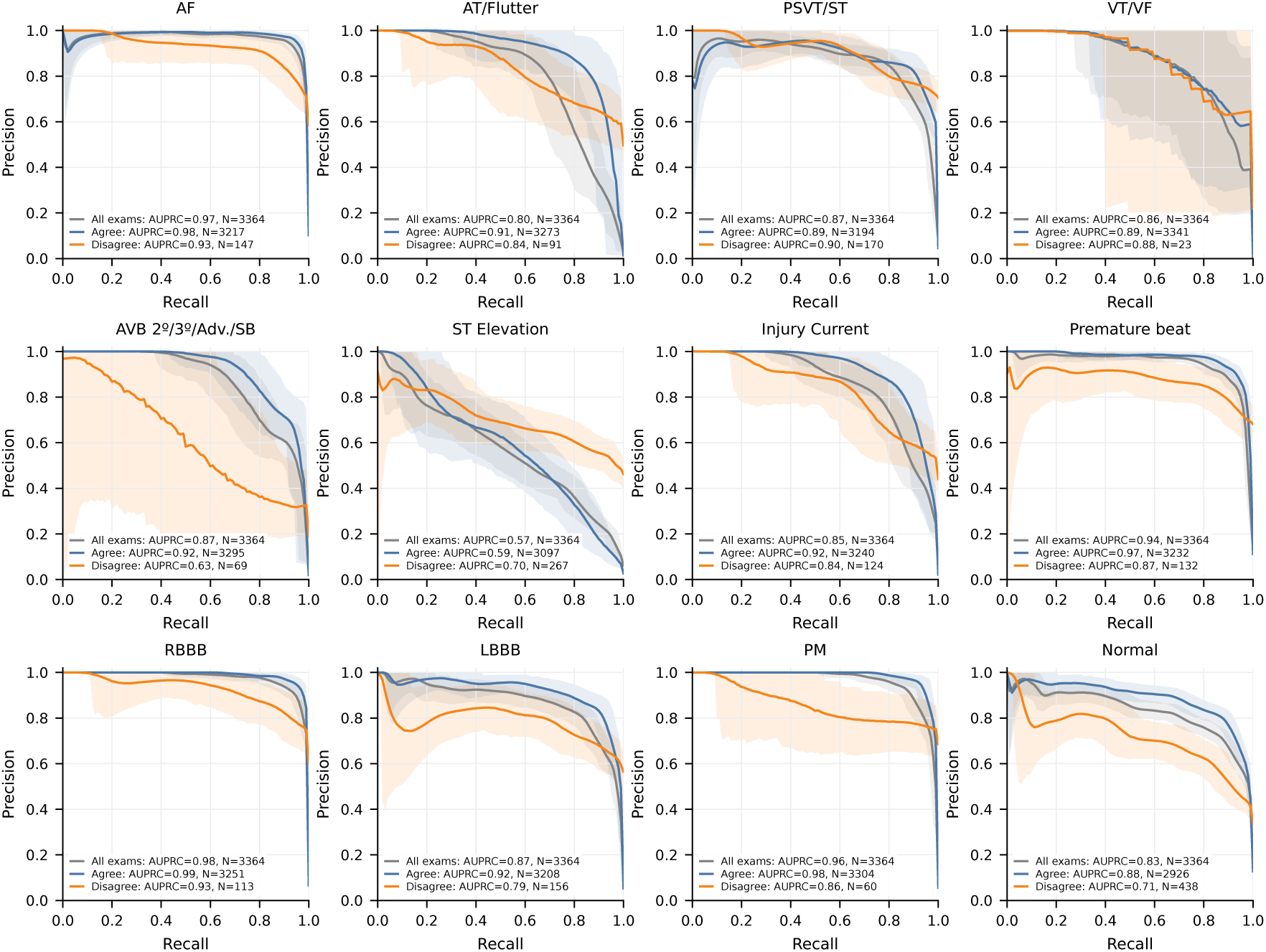
Class-wise precision-recall curves stratified by primary-reader agreement. Curves are shown separately for all held-out test ECGs, ECGs in which the two primary cardiologist readers agreed and ECGs in which they disagreed before adjudication.

**Supplementary Figure 10.**
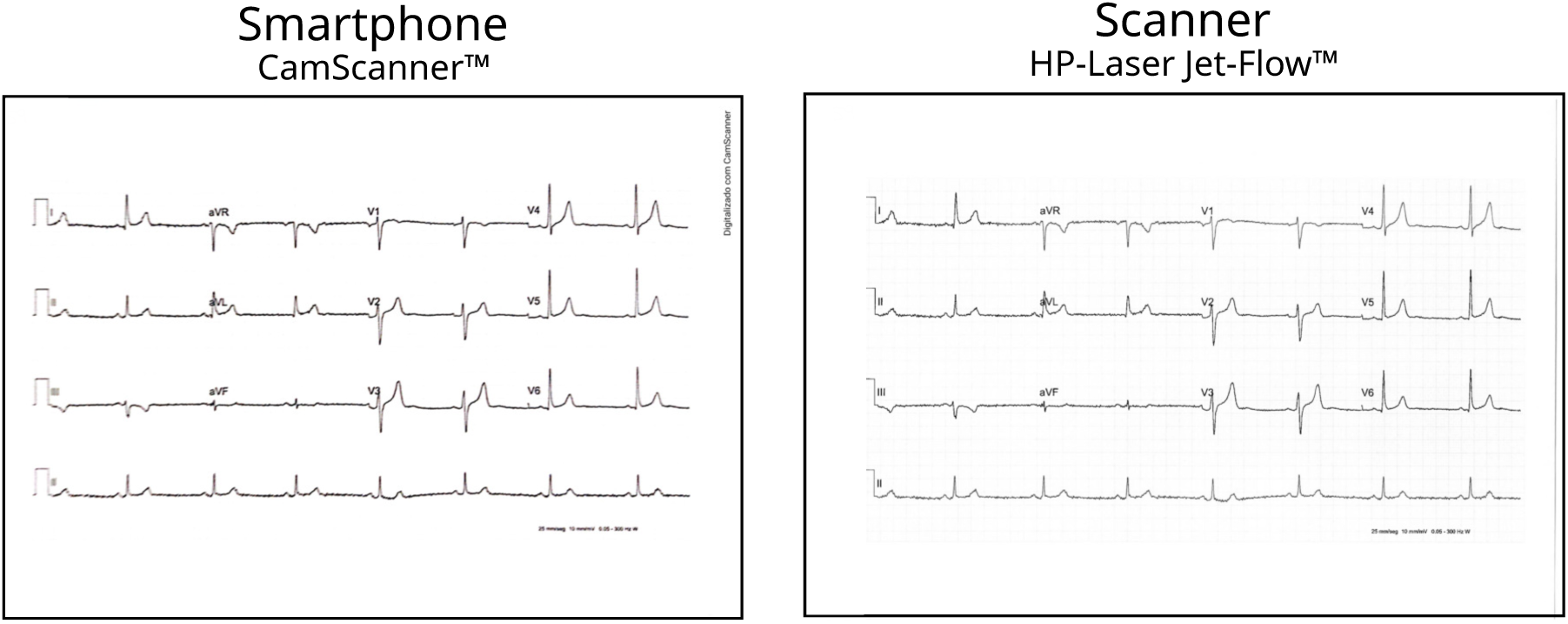
Example of scanned and photographed ECG image used in the acquisition-shift evaluation.

**Supplementary Figure 11.**
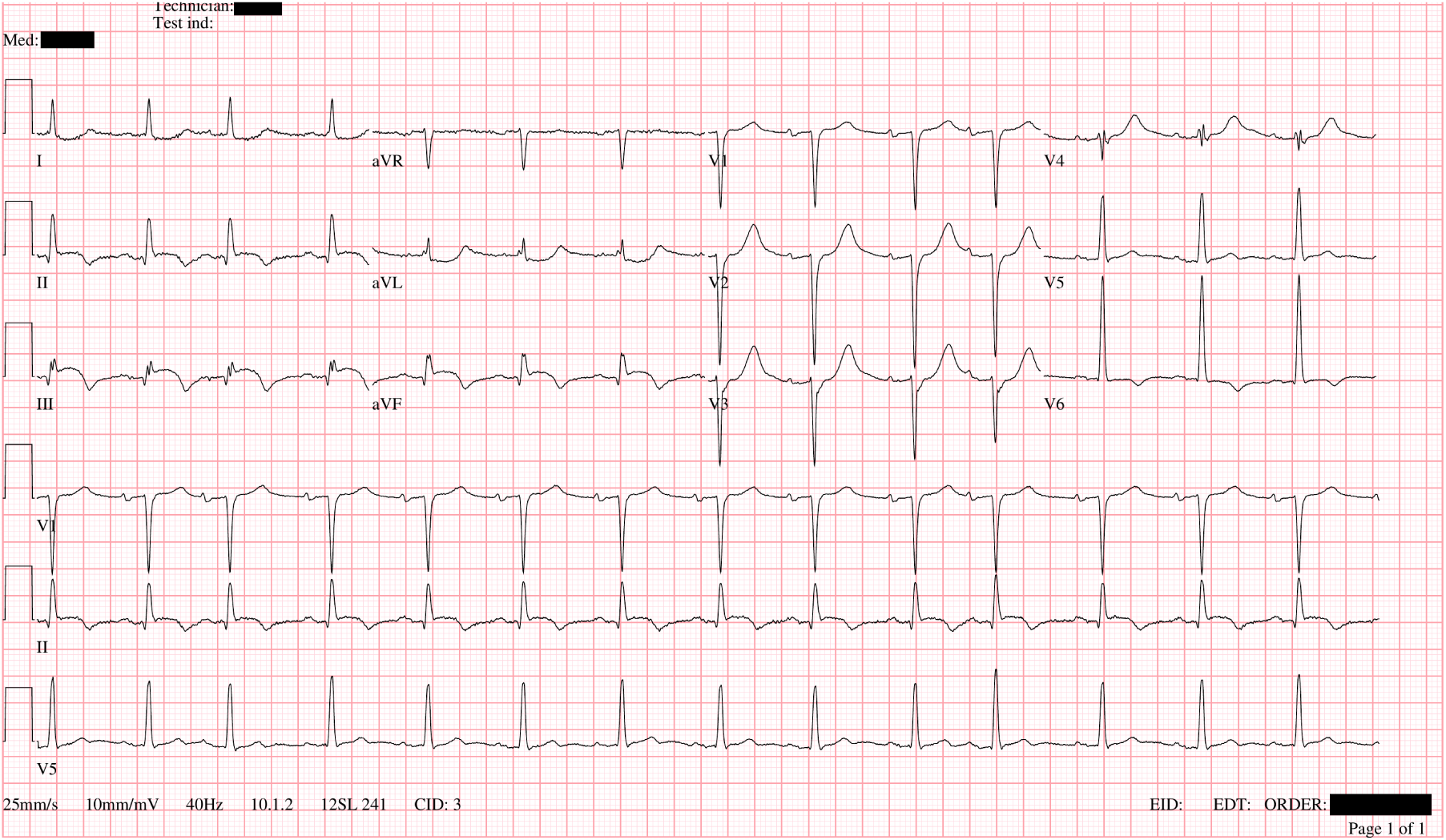
Representative ECG image from the enriched temporal emergency evaluation. These future emergency ECGs were acquired after the InCor-EMG study period using a GE-MUSE system and had an image format different from the Mortara ELI 250c images used in InCor-EMG.

## Supplementary Appendix B Supplementary Tables

**Supplementary Table 1.**
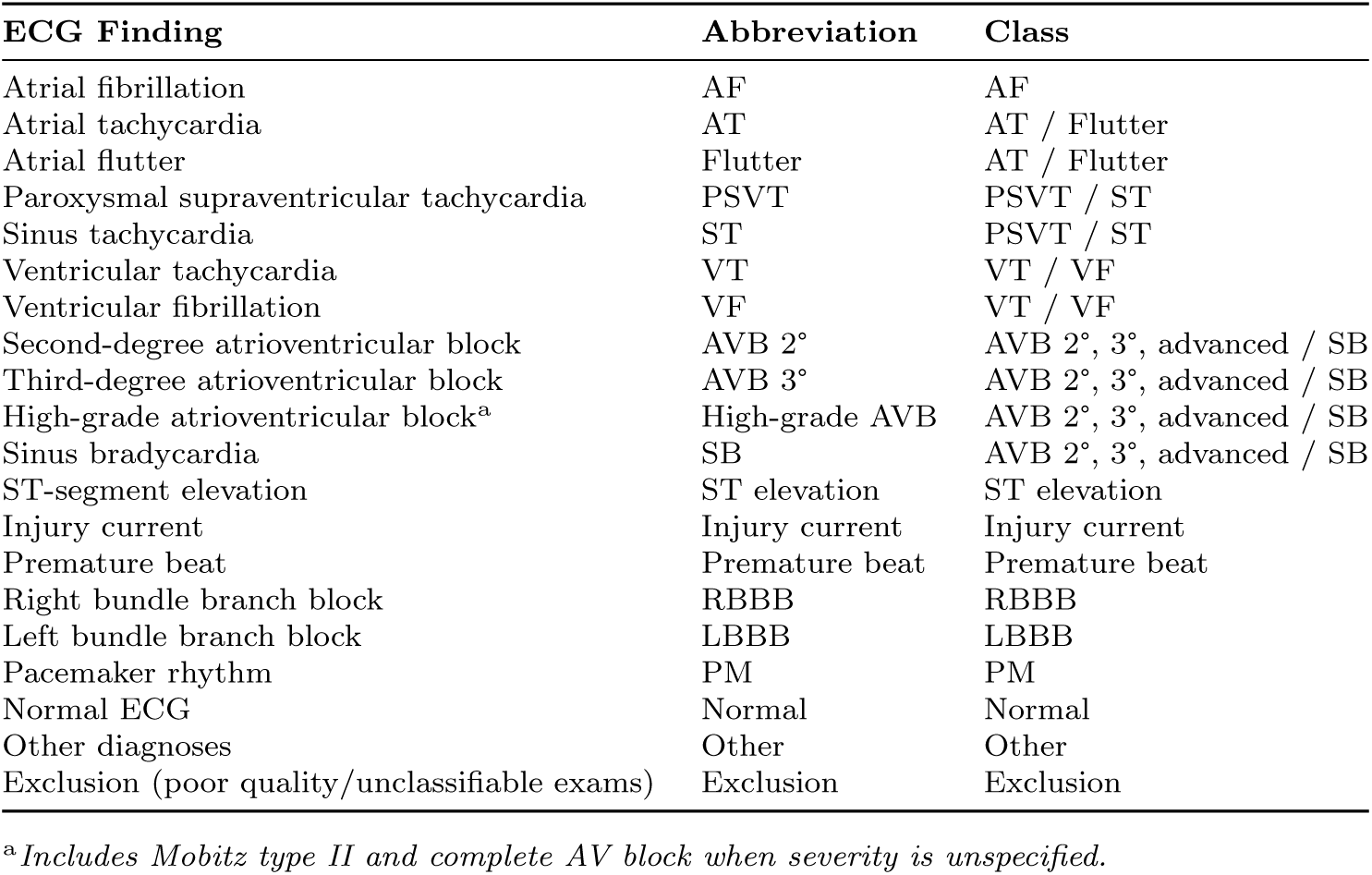
Emergency ECG classes and modeled ECG category used in InCor-EMG. Candidate ECG findings were grouped into clinically related classes for multi-label model training and evaluation.

**Supplementary Table 2.**
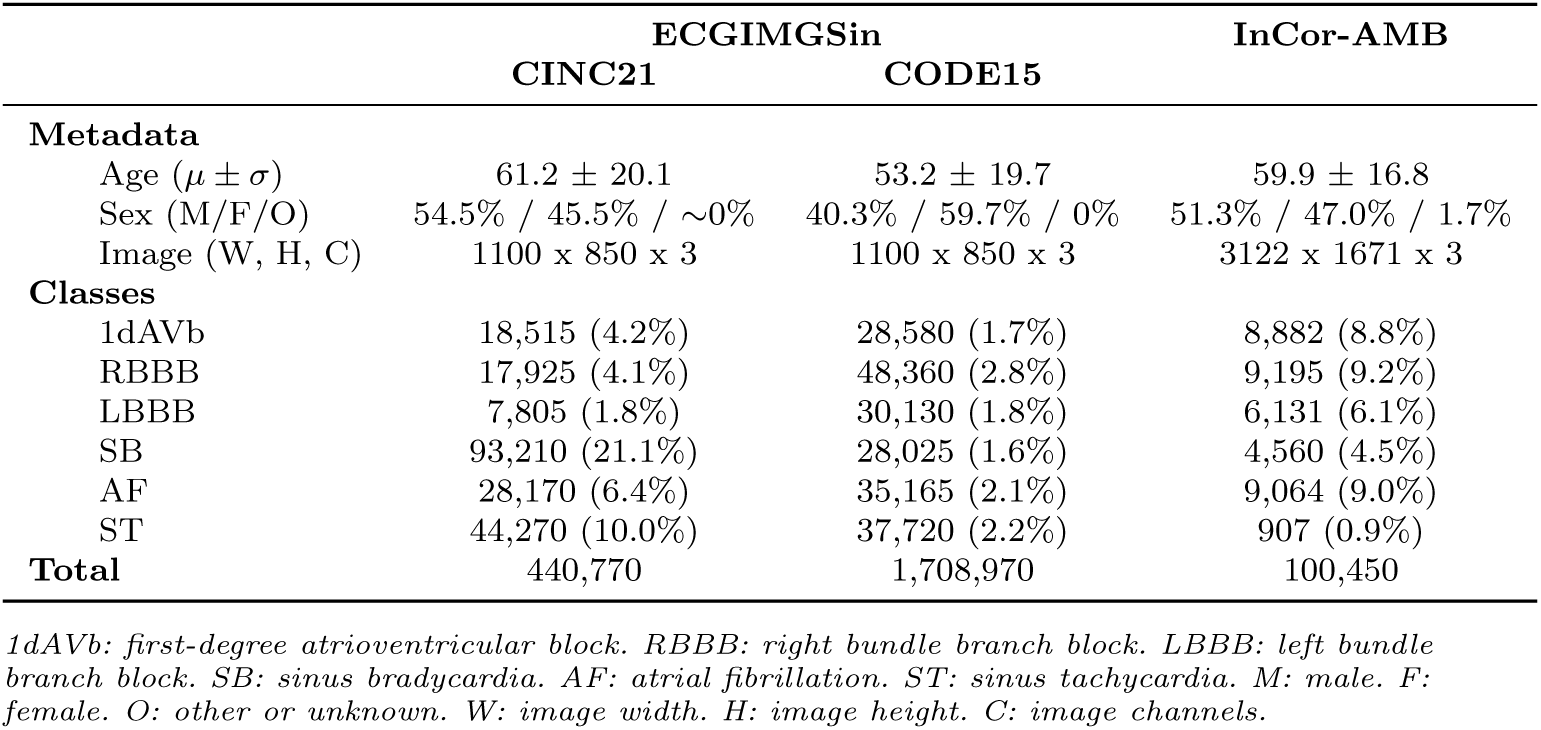
Summary of the ECGIMGSin and InCor-AMB datasets used for pretraining and intermediate model development. ECGIMGSin was generated from CODE15 and CINC21 waveforms rendered as synthetic ECG images, whereas InCor-AMB contained real ambulatory ECG images extracted from institutional DICOM files. Percentages are computed within each dataset.

**Supplementary Table 3.** Mean F1-score obtained by the candidate architectures on the 10-fold cross-validation performed within the InCor-EMG training split. No training optimization was included in these architecture-selection runs.

| Classes | ConvNeXt | EfficientNet-B3 | ResNet50 | ViT | SwinT |
| --- | --- | --- | --- | --- | --- |
| AF | **0.875** | 0.808 | 0.833 | 0.737 | 0.810 |
| AT/Flutter | **0.572** | 0.565 | 0.549 | 0.512 | 0.568 |
| PSVT/ST | **0.754** | 0.752 | 0.753 | 0.694 | 0.727 |
| VT/VF | **0.678** | 0.603 | 0.659 | 0.549 | 0.597 |
| AVB 2nd/3rd/advanced/SB | 0.687 | 0.684 | **0.694** | 0.585 | 0.681 |
| ST elevation | **0.479** | 0.426 | 0.411 | 0.400 | 0.412 |
| Injury current | **0.725** | 0.695 | 0.706 | 0.553 | 0.643 |
| Premature beats | **0.844** | 0.802 | 0.802 | 0.645 | 0.775 |
| RBBB | **0.874** | 0.843 | 0.848 | 0.816 | 0.863 |
| LBBB | **0.806** | 0.784 | 0.787 | 0.761 | 0.792 |
| PM | **0.847** | 0.794 | 0.816 | 0.806 | 0.843 |
| Normal | 0.699 | 0.694 | **0.706** | 0.678 | 0.697 |
| **Mean** | **0.737** | 0.704 | 0.714 | 0.645 | 0.701 |

**Supplementary Table 4.**
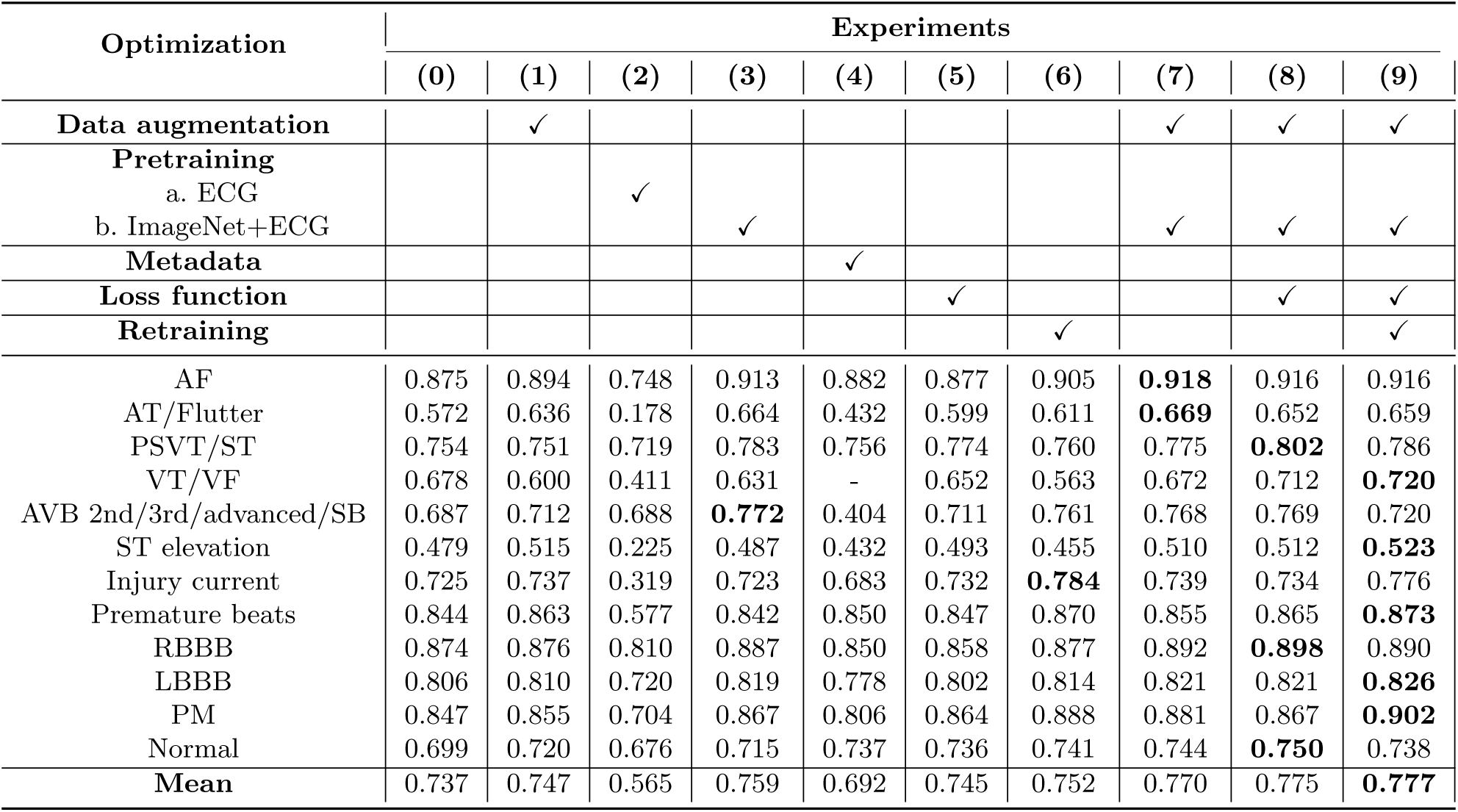
Ablation study reporting mean F1-score across 10-fold cross-validation for the proposed performance-optimization strategies. Values were reconstructed from the available class- and fold-level confusion-matrix CSV files; the source run for each column is documented in training optimization 10col source runs.csv.

**Supplementary Table 5.** Internal test performance of the final 10-fold ConvNeXt ensemble on the InCor-EMG test set. Binary predictions used a fixed threshold of 0.5 for all classes. Confidence intervals were computed with 1,000 patient-level cluster bootstrap resamples.

| Class | AUROC (95% CI) | AUPRC (95% CI) | Se (95% CI) | Spe (95% CI) | PPV (95% CI) | F1 (95% CI) |
| --- | --- | --- | --- | --- | --- | --- |
| AF (N=414) | 0.997 (0.995-0.998) | 0.970 (0.947-0.986) | 0.937 (0.913-0.960) | 0.990 (0.986-0.994) | 0.930 (0.903-0.955) | 0.934 (0.914-0.950) |
| AT/Flutter (N=90) | 0.978 (0.959-0.991) | 0.800 (0.717-0.874) | 0.633 (0.526-0.735) | 0.998 (0.996-0.999) | 0.891 (0.808-0.962) | 0.740 (0.653-0.818) |
| PSVT/ST (N=266) | 0.983 (0.975-0.989) | 0.868 (0.821-0.909) | 0.789 (0.741-0.838) | 0.989 (0.985-0.993) | 0.861 (0.815-0.904) | 0.824 (0.784-0.856) |
| VT/VF (N=17) | 0.999 (0.997-1.000) | 0.863 (0.698-0.976) | 0.765 (0.571-0.945) | 0.999 (0.998-1.000) | 0.812 (0.600-1.000) | 0.788 (0.621-0.923) |
| AVB 2°/3°/Adv./SB (N=76) | 0.993 (0.986-0.998) | 0.875 (0.805-0.928) | 0.789 (0.683-0.875) | 0.994 (0.991-0.996) | 0.741 (0.640-0.830) | 0.764 (0.677-0.831) |
| ST Elevation (N=207) | 0.933 (0.914-0.948) | 0.573 (0.498-0.641) | 0.507 (0.437-0.570) | 0.977 (0.972-0.983) | 0.593 (0.514-0.665) | 0.547 (0.483-0.605) |
| Injury Current (N=124) | 0.989 (0.983-0.994) | 0.854 (0.791-0.902) | 0.774 (0.688-0.852) | 0.993 (0.990-0.996) | 0.807 (0.737-0.874) | 0.790 (0.723-0.845) |
| Premature beat (N=451) | 0.986 (0.978-0.991) | 0.943 (0.921-0.962) | 0.882 (0.852-0.914) | 0.986 (0.981-0.990) | 0.905 (0.878-0.931) | 0.893 (0.871-0.915) |
| RBBB (N=283) | 0.998 (0.997-0.999) | 0.978 (0.965-0.987) | 0.943 (0.910-0.971) | 0.992 (0.988-0.995) | 0.911 (0.879-0.942) | 0.927 (0.902-0.949) |
| LBBB (N=256) | 0.985 (0.978-0.991) | 0.869 (0.820-0.910) | 0.855 (0.810-0.897) | 0.982 (0.976-0.987) | 0.796 (0.742-0.845) | 0.825 (0.787-0.859) |
| PM (N=225) | 0.996 (0.993-0.998) | 0.962 (0.941-0.978) | 0.800 (0.744-0.857) | 0.998 (0.996-0.999) | 0.963 (0.934-0.986) | 0.874 (0.835-0.909) |
| Normal (N=521) | 0.969 (0.963-0.974) | 0.830 (0.793-0.862) | 0.812 (0.778-0.845) | 0.952 (0.944-0.960) | 0.757 (0.721-0.793) | 0.783 (0.756-0.812) |
| Macro mean | 0.984 (0.981-0.986) | 0.865 (0.847-0.882) | 0.791 (0.767-0.815) | 0.987 (0.986-0.989) | 0.831 (0.806-0.852) | 0.807 (0.788-0.825) |

**Supplementary Table 6.**
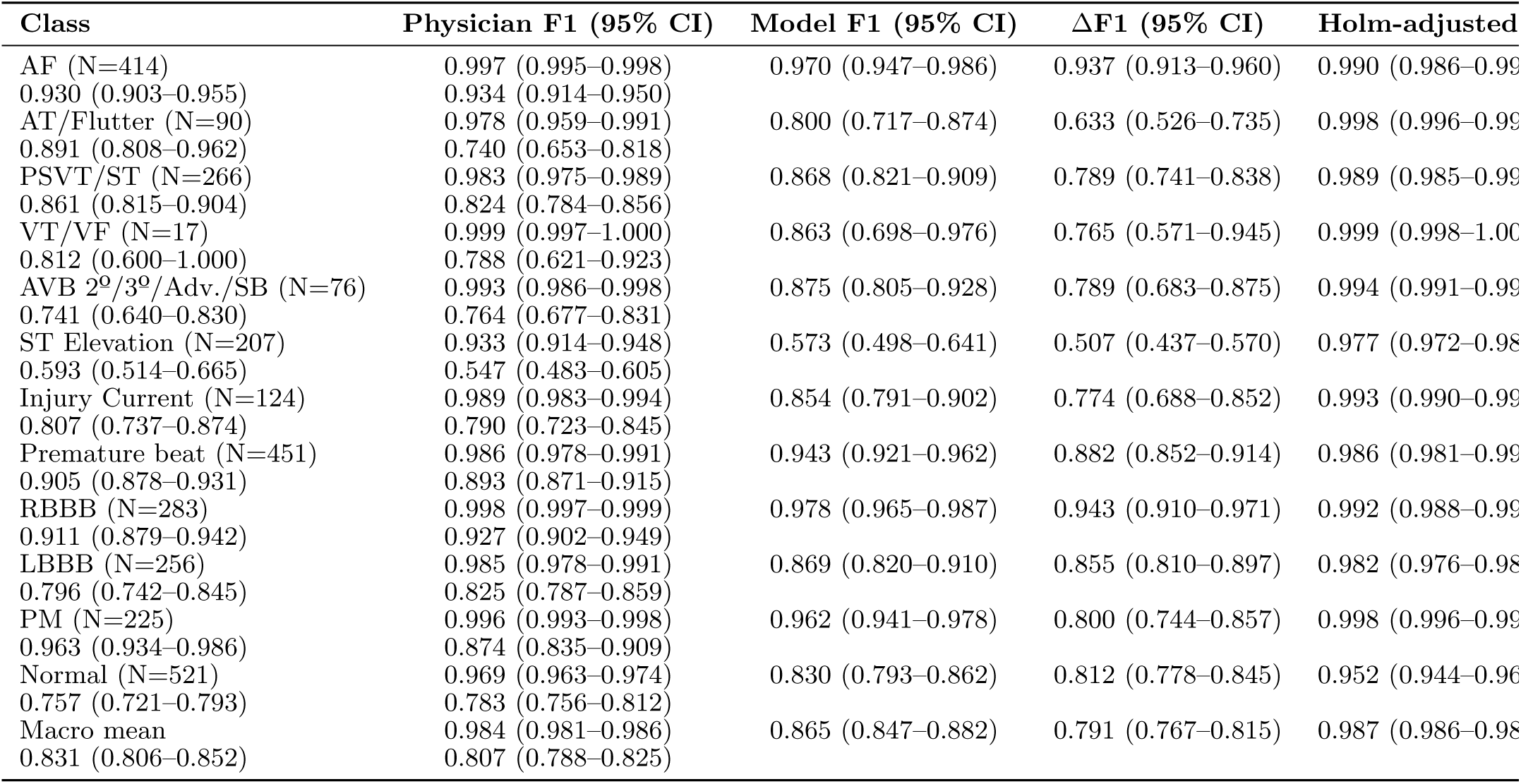
Class-wise comparison between physicians and the final model on the same InCor-EMG internal test ECGs. T analysis used both independent reader instances when available. Binary model predictions used a fixed threshold of 0.5 for all classes. Confidence intervals were computed with 1,000 patient-level cluster bootstrap resamples. P values used paired patient-level permutation a were Holm-adjusted across classes. The macro mean row was computed by averaging class-wise bootstrap draws; its p value is a two-sided bootstrap p value for the macro delta and was not included in class-wise multiplicity correction.

**Supplementary Table 7.** Class-wise comparison between Mortara Veritas^™^ and the final model on the same InCor-EMG internal test ECGs. Binary model predictions used a fixed threshold of 0.5 for all classes; Mortara labels used the prespecified statement mapping. Confidence intervals were computed with 1,000 paired patient-level cluster bootstrap resamples. P values used paired patient-level permutation and were Holm-adjusted across classes. The macro mean row was computed by averaging class-wise bootstrap draws; its p value is a two-sided bootstrap p value for the macro delta and was not included in class-wise multiplicity correction.

| Class | Mortara F1 (95% CI) | Model F1 (95% CI) | $\Delta$ F1 (95% CI) | Holm-adjusted p |
| --- | --- | --- | --- | --- |
| AF | 0.843 (0.814–0.871) | 0.934 (0.915–0.951) | 0.091 (0.063–0.119) | 0.012 |
| AT/Flutter | 0.452 (0.342–0.553) | 0.740 (0.646–0.821) | 0.288 (0.153–0.417) | 0.012 |
| PSVT/ST | 0.397 (0.354–0.439) | 0.824 (0.784–0.859) | 0.426 (0.380–0.470) | 0.012 |
| VT/VF | 0.000 (0.000–0.000) | 0.788 (0.609–0.909) | 0.788 (0.609–0.909) | 0.012 |
| AVB 2<sup>o</sup>/3<sup>o</sup>/Adv./SB | 0.142 (0.106–0.179) | 0.764 (0.678–0.835) | 0.623 (0.546–0.693) | 0.012 |
| ST Elevation | 0.158 (0.097–0.221) | 0.547 (0.487–0.604) | 0.389 (0.316–0.461) | 0.012 |
| Injury Current | 0.174 (0.143–0.206) | 0.790 (0.728–0.844) | 0.616 (0.560–0.670) | 0.012 |
| Premature beat | 0.746 (0.711–0.777) | 0.893 (0.870–0.915) | 0.148 (0.116–0.182) | 0.012 |
| RBBB | 0.719 (0.670–0.759) | 0.927 (0.903–0.949) | 0.208 (0.166–0.254) | 0.012 |
| LBBB | 0.689 (0.632–0.741) | 0.825 (0.786–0.860) | 0.136 (0.083–0.191) | 0.012 |
| PM | 0.846 (0.802–0.884) | 0.874 (0.834–0.909) | 0.028 (-0.022–0.079) | 0.308 |
| Normal | 0.000 (0.000–0.000) | 0.783 (0.755–0.809) | 0.783 (0.755–0.809) | 0.012 |
| Macro mean | 0.431 (0.416–0.444) | 0.806 (0.786–0.824) | 0.376 (0.353–0.397) | <0.001 |

**Supplementary Table 8.**
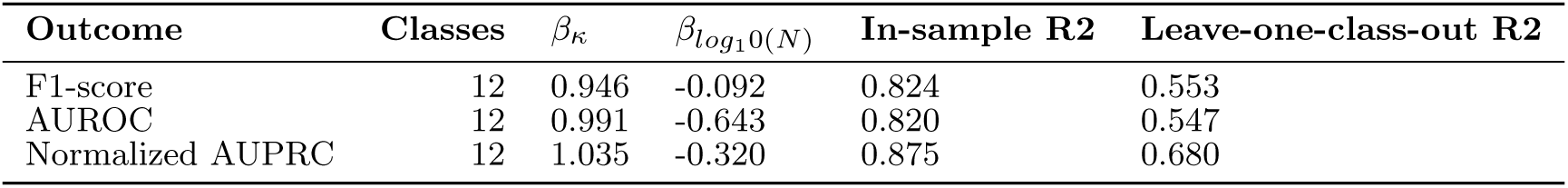
Standardized multivariable class-level models evaluating determinants of model performance. Each model included inter-reader kappa and the log-transformed number of positive training examples. These ecological class-level analyses support association, not causality.

| Outcome | Classes | $\beta_\kappa$ | $\beta_{\log_{10}(N)}$ | In-sample R2 | Leave-one-class-out R2 |
| --- | --- | --- | --- | --- | --- |
| F1-score | 12 | 0.946 | -0.092 | 0.824 | 0.553 |
| AUROC | 12 | 0.991 | -0.643 | 0.820 | 0.547 |
| Normalized AUPRC | 12 | 1.035 | -0.320 | 0.875 | 0.680 |

**Supplementary Table 9.** Class-wise model performance under acquisition shift. AUROC and AUPRC were computed from continuous model probabilities for each acquisition condition. Confidence intervals were computed with 1,000 ECG-level bootstrap resamples.

| Class | Scenario | AUROC (95% CI) | AUPRC (95% CI) | Se (95% CI) | Spe (95% CI) | PPV (95% CI) | F1 (95% CI) |
| --- | --- | --- | --- | --- | --- | --- | --- |
| AF (N=17) | Original | 0.990 (0.973–1.000) | 0.939 (0.820–1.000) | 0.941 (0.812–1.000) | 0.971 (0.935–1.000) | 0.842 (0.667–1.000) | 0.889 (0.758–0.980) |
|  | Scan | 0.990 (0.971–1.000) | 0.926 (0.771–1.000) | 0.941 (0.809–1.000) | 0.951 (0.906–0.990) | 0.762 (0.555–0.944) | 0.842 (0.688–0.947) |
|  | Photo | 0.987 (0.964–1.000) | 0.910 (0.741–1.000) | 0.941 (0.786–1.000) | 0.961 (0.922–0.991) | 0.800 (0.600–0.955) | 0.865 (0.714–0.966) |
| AT/Flutter (N=14) | Original | 0.946 (0.866–0.995) | 0.832 (0.644–0.969) | 0.571 (0.333–0.846) | 1.000 (1.000–1.000) | 1.000 (1.000–1.000) | 0.727 (0.500–0.917) |
|  | Scan | 0.942 (0.873–0.996) | 0.831 (0.642–0.976) | 0.286 (0.071–0.556) | 1.000 (1.000–1.000) | 1.000 (1.000–1.000) | 0.444 (0.133–0.714) |
|  | Photo | 0.947 (0.884–0.995) | 0.836 (0.640–0.969) | 0.429 (0.154–0.700) | 1.000 (1.000–1.000) | 1.000 (1.000–1.000) | 0.600 (0.267–0.824) |
| PSVT/ST (N=13) | Original | 0.955 (0.886–0.995) | 0.835 (0.604–0.966) | 0.692 (0.400–0.917) | 0.972 (0.937–1.000) | 0.750 (0.500–1.000) | 0.720 (0.454–0.889) |
|  | Scan | 0.909 (0.736–0.996) | 0.779 (0.516–0.978) | 0.769 (0.500–1.000) | 0.963 (0.925–0.991) | 0.714 (0.462–0.929) | 0.741 (0.500–0.903) |
|  | Photo | 0.912 (0.741–0.996) | 0.785 (0.508–0.973) | 0.769 (0.500–1.000) | 0.981 (0.952–1.000) | 0.833 (0.556–1.000) | 0.800 (0.571–0.938) |
| VT/VF (N=10) | Original | 0.996 (0.987–1.000) | 0.965 (0.859–1.000) | 0.600 (0.286–0.909) | 1.000 (1.000–1.000) | 1.000 (1.000–1.000) | 0.750 (0.444–0.952) |
|  | Scan | 0.995 (0.983–1.000) | 0.950 (0.795–1.000) | 0.500 (0.167–0.833) | 1.000 (1.000–1.000) | 1.000 (1.000–1.000) | 0.667 (0.286–0.909) |
|  | Photo | 0.993 (0.978–1.000) | 0.923 (0.732–1.000) | 0.500 (0.200–0.833) | 1.000 (1.000–1.000) | 1.000 (1.000–1.000) | 0.667 (0.333–0.909) |
| AVB 2°/3°/Adv./SB (N=12) | Original | 0.998 (0.992–1.000) | 0.987 (0.933–1.000) | 0.833 (0.600–1.000) | 1.000 (1.000–1.000) | 1.000 (1.000–1.000) | 0.909 (0.750–1.000) |
|  | Scan | 0.994 (0.979–1.000) | 0.959 (0.858–1.000) | 0.667 (0.375–0.923) | 1.000 (1.000–1.000) | 1.000 (1.000–1.000) | 0.800 (0.545–0.960) |
|  | Photo | 0.995 (0.982–1.000) | 0.963 (0.870–1.000) | 0.750 (0.500–1.000) | 1.000 (1.000–1.000) | 1.000 (1.000–1.000) | 0.857 (0.667–1.000) |
| ST Elevation (N=11) | Original | 0.950 (0.861–0.997) | 0.785 (0.523–0.968) | 0.455 (0.167–0.800) | 0.991 (0.971–1.000) | 0.833 (0.500–1.000) | 0.588 (0.267–0.857) |
|  | Scan | 0.958 (0.916–0.990) | 0.642 (0.346–0.899) | 0.364 (0.083–0.700) | 0.982 (0.955–1.000) | 0.667 (0.200–1.000) | 0.471 (0.143–0.762) |
|  | Photo | 0.945 (0.885–0.990) | 0.644 (0.360–0.919) | 0.364 (0.083–0.667) | 0.982 (0.954–1.000) | 0.667 (0.200–1.000) | 0.471 (0.133–0.727) |
| Injury Current (N=11) | Original | 0.999 (0.995–1.000) | 0.992 (0.954–1.000) | 0.818 (0.556–1.000) | 1.000 (1.000–1.000) | 1.000 (1.000–1.000) | 0.900 (0.714–1.000) |
|  | Scan | 0.998 (0.992–1.000) | 0.984 (0.913–1.000) | 0.727 (0.444–1.000) | 1.000 (1.000–1.000) | 1.000 (1.000–1.000) | 0.842 (0.615–1.000) |
|  | Photo | 0.997 (0.990–1.000) | 0.975 (0.880–1.000) | 0.727 (0.429–1.000) | 1.000 (1.000–1.000) | 1.000 (1.000–1.000) | 0.842 (0.600–1.000) |
| Premature beat (N=20) | Original | 0.995 (0.984–1.000) | 0.979 (0.926–1.000) | 0.900 (0.737–1.000) | 0.980 (0.947–1.000) | 0.900 (0.733–1.000) | 0.900 (0.774–0.978) |
|  | Scan | 0.953 (0.843–1.000) | 0.913 (0.760–1.000) | 0.850 (0.684–1.000) | 0.990 (0.968–1.000) | 0.944 (0.800–1.000) | 0.895 (0.759–0.980) |
|  | Photo | 0.960 (0.878–1.000) | 0.932 (0.825–0.998) | 0.850 (0.667–1.000) | 0.990 (0.967–1.000) | 0.944 (0.812–1.000) | 0.895 (0.769–0.977) |
| RBBB (N=14) | Original | 1.000 (1.000–1.000) | 1.000 (1.000–1.000) | 1.000 (1.000–1.000) | 1.000 (1.000–1.000) | 1.000 (1.000–1.000) | 1.000 (1.000–1.000) |
|  | Scan | 1.000 (1.000–1.000) | 1.000 (1.000–1.000) | 1.000 (1.000–1.000) | 1.000 (1.000–1.000) | 1.000 (1.000–1.000) | 1.000 (1.000–1.000) |
|  | Photo | 1.000 (1.000–1.000) | 1.000 (1.000–1.000) | 0.929 (0.769–1.000) | 1.000 (1.000–1.000) | 1.000 (1.000–1.000) | 0.963 (0.869–1.000) |
| LBBB (N=15) | Original | 0.950 (0.899–0.985) | 0.760 (0.531–0.915) | 0.800 (0.571–1.000) | 0.943 (0.897–0.981) | 0.667 (0.438–0.875) | 0.727 (0.522–0.875) |
|  | Scan | 0.964 (0.925–0.992) | 0.807 (0.592–0.950) | 0.800 (0.571–1.000) | 0.933 (0.880–0.980) | 0.632 (0.400–0.850) | 0.706 (0.483–0.864) |
|  | Photo | 0.971 (0.941–0.993) | 0.835 (0.651–0.962) | 0.800 (0.583–1.000) | 0.943 (0.896–0.981) | 0.667 (0.438–0.882) | 0.727 (0.529–0.878) |
| PM (N=11) | Original | 0.995 (0.983–1.000) | 0.960 (0.845–1.000) | 0.818 (0.545–1.000) | 0.982 (0.953–1.000) | 0.818 (0.571–1.000) | 0.818 (0.600–0.963) |
|  | Scan | 0.989 (0.968–1.000) | 0.924 (0.773–1.000) | 0.818 (0.556–1.000) | 0.982 (0.952–1.000) | 0.818 (0.538–1.000) | 0.818 (0.588–0.957) |
|  | Photo | 0.991 (0.972–1.000) | 0.929 (0.753–1.000) | 0.818 (0.545–1.000) | 0.972 (0.936–1.000) | 0.750 (0.455–1.000) | 0.783 (0.526–0.941) |
| Normal (N=10) | Original | 0.995 (0.980–1.000) | 0.963 (0.842–1.000) | 0.900 (0.667–1.000) | 1.000 (1.000–1.000) | 1.000 (1.000–1.000) | 0.947 (0.800–1.000) |
|  | Scan | 0.991 (0.971–1.000) | 0.934 (0.779–1.000) | 0.800 (0.500–1.000) | 0.982 (0.954–1.000) | 0.800 (0.500–1.000) | 0.800 (0.545–0.957) |
|  | Photo | 0.995 (0.977–1.000) | 0.963 (0.837–1.000) | 0.900 (0.667–1.000) | 0.973 (0.936–1.000) | 0.750 (0.444–1.000) | 0.818 (0.571–0.960) |
| Macro mean | Original | 0.981 (0.970–0.990) | 0.916 (0.876–0.949) | 0.777 (0.717–0.842) | 0.987 (0.980–0.993) | 0.901 (0.852–0.947) | 0.823 (0.763–0.865) |
|  | Scan | 0.974 (0.955–0.986) | 0.888 (0.838–0.929) | 0.710 (0.637–0.778) | 0.982 (0.974–0.989) | 0.861 (0.803–0.914) | 0.752 (0.670–0.802) |
|  | Photo | 0.974 (0.956–0.987) | 0.891 (0.845–0.931) | 0.731 (0.661–0.798) | 0.984 (0.975–0.990) | 0.868 (0.806–0.920) | 0.774 (0.702–0.823) |

**Supplementary Table 10.** Pairwise F1-score differences relative to original digital ECG images in the 120-ECG acquisition-shift subset. Positive delta values indicate higher F1 for original digital images. Confidence intervals were computed with 1,000 paired ECG-level bootstrap resamples; p values were adjusted for multiple testing using the Holm method.

| Class | Original - Scan $\Delta$ F1 (95% CI) | Holm-adjusted p | Original - Photo $\Delta$ F1 (95% CI) | Holm-adjusted p |
| --- | --- | --- | --- | --- |
| AF (N=17) | 0.047 (0.000–0.131) | 1.000 | 0.024 (-0.058–0.124) | 1.000 |
| AT/Flutter (N=14) | 0.283 (0.063–0.583) | 0.624 | 0.127 (0.000–0.338) | 1.000 |
| PSVT/ST (N=13) | -0.021 (-0.171–0.083) | 1.000 | -0.080 (-0.229–0.000) | 1.000 |
| VT/VF (N=10) | 0.083 (0.000–0.318) | 1.000 | 0.083 (0.000–0.318) | 1.000 |
| AVB 2°/3°/Adv./SB (N=12) | 0.109 (0.000–0.309) | 1.000 | 0.052 (0.000–0.197) | 1.000 |
| ST Elevation (N=11) | 0.118 (0.000–0.357) | 1.000 | 0.118 (0.000–0.341) | 1.000 |
| Injury Current (N=11) | 0.058 (0.000–0.227) | 1.000 | 0.058 (0.000–0.227) | 1.000 |
| Premature beat (N=20) | 0.005 (-0.067–0.094) | 1.000 | 0.005 (-0.067–0.094) | 1.000 |
| RBBB (N=14) | 0.000 (0.000–0.000) | 1.000 | 0.037 (0.000–0.130) | 1.000 |
| LBBB (N=15) | 0.021 (0.000–0.070) | 1.000 | 0.000 (0.000–0.000) | 1.000 |
| PM (N=11) | 0.000 (0.000–0.000) | 1.000 | 0.036 (0.000–0.129) | 1.000 |
| Normal (N=10) | 0.147 (0.000–0.376) | 1.000 | 0.129 (0.000–0.321) | 1.000 |

**Supplementary Table 11.**
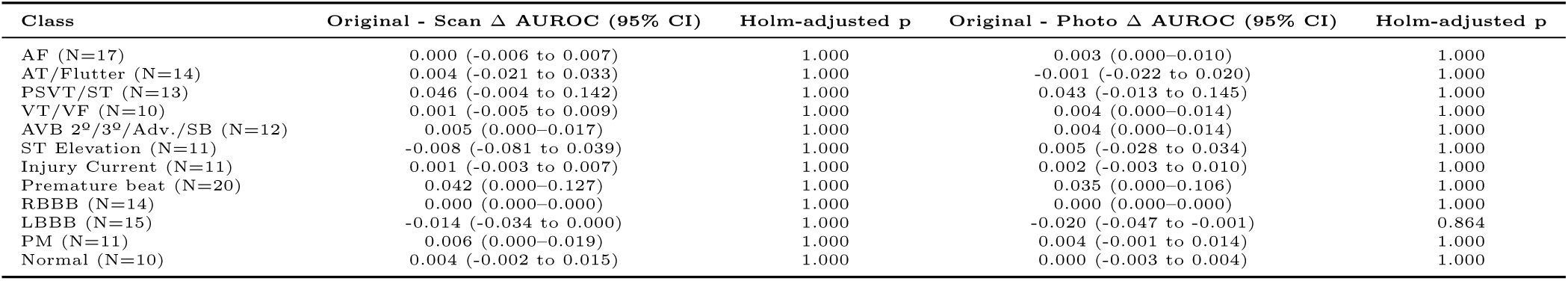
Pairwise AUROC differences relative to original digital ECG images in the 120-ECG acquisition-shift subset. Positive delta values indicate higher AUROC for original digital images. Confidence intervals were computed with 1,000 ECG-level bootstrap resamples; p values were adjusted for multiple testing using the Holm method.

| Class | Original - Scan $\Delta$ AUROC (95% CI) | Holm-adjusted p | Original - Photo $\Delta$ AUROC (95% CI) | Holm-adjusted p |
| --- | --- | --- | --- | --- |
| AF (N=17) | 0.000 (-0.006 to 0.007) | 1.000 | 0.003 (0.000–0.010) | 1.000 |
| AT/Flutter (N=14) | 0.004 (-0.021 to 0.033) | 1.000 | -0.001 (-0.022 to 0.020) | 1.000 |
| PSVT/ST (N=13) | 0.046 (-0.004 to 0.142) | 1.000 | 0.043 (-0.013 to 0.145) | 1.000 |
| VT/VF (N=10) | 0.001 (-0.005 to 0.009) | 1.000 | 0.004 (0.000–0.014) | 1.000 |
| AVB 2°/3°/Adv./SB (N=12) | 0.005 (0.000–0.017) | 1.000 | 0.004 (0.000–0.014) | 1.000 |
| ST Elevation (N=11) | -0.008 (-0.081 to 0.039) | 1.000 | 0.005 (-0.028 to 0.034) | 1.000 |
| Injury Current (N=11) | 0.001 (-0.003 to 0.007) | 1.000 | 0.002 (-0.003 to 0.010) | 1.000 |
| Premature beat (N=20) | 0.042 (0.000–0.127) | 1.000 | 0.035 (0.000–0.106) | 1.000 |
| RBBB (N=14) | 0.000 (0.000–0.000) | 1.000 | 0.000 (0.000–0.000) | 1.000 |
| LBBB (N=15) | -0.014 (-0.034 to 0.000) | 1.000 | -0.020 (-0.047 to -0.001) | 0.864 |
| PM (N=11) | 0.006 (0.000–0.019) | 1.000 | 0.004 (-0.001 to 0.014) | 1.000 |
| Normal (N=10) | 0.004 (-0.002 to 0.015) | 1.000 | 0.000 (-0.003 to 0.004) | 1.000 |

**Supplementary Table 12.**
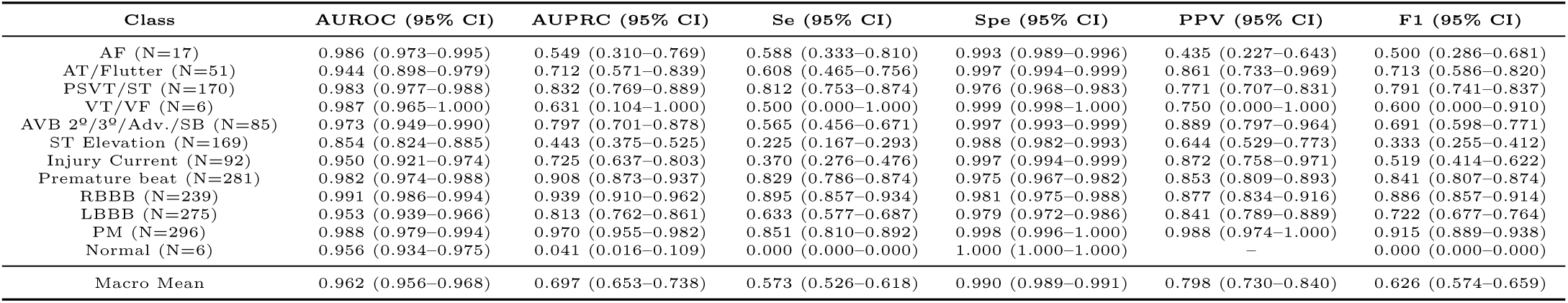
Class-wise performance of the final 10-fold ConvNeXt ensemble in the enriched temporal emergency evaluation. Class counts indicate the number of positive labels in the multi-label reference standard. Confidence intervals were computed with 1,000 patient-level cluster bootstrap resamples.

**Supplementary Table 13.** Class-wise AUROC differences between the enriched temporal emergency evaluation and the final InCor-EMG internal test set. Delta values are enriched temporal evaluation minus internal test. Confidence intervals and p values were obtained from 1,000 bootstrap metric draws; p values were adjusted across classes using the Holm method.

| Class | Temporal - Internal $\Delta$ AUROC (95% CI) | Holm-adjusted p |
| --- | --- | --- |
| AF | -0.011 (-0.025 to -0.001) | 0.196 |
| AT/Flutter | -0.034 (-0.082 to 0.004) | 0.530 |
| PSVT/ST | 0.000 (-0.009 to 0.009) | 0.984 |
| VT/VF | -0.013 (-0.033 to 0.002) | 0.826 |
| AVB 2<sup>o</sup>/3<sup>o</sup>/Adv./SB | -0.021 (-0.045 to -0.002) | 0.196 |
| ST Elevation | -0.078 (-0.113 to -0.041) | 0.000 |
| Injury Current | -0.039 (-0.068 to -0.016) | 0.000 |
| Premature beat | -0.004 (-0.014 to 0.006) | 0.884 |
| RBBB | -0.007 (-0.012 to -0.003) | 0.000 |
| LBBB | -0.032 (-0.048 to -0.018) | 0.000 |
| PM | -0.008 (-0.017 to -0.001) | 0.176 |
| Normal | -0.013 (-0.036 to 0.008) | 0.826 |

**Supplementary Table 14.** Class-wise performance of the final 10-fold ConvNeXt ensemble on the external Life in the Fast Lane ECG image set. Class counts indicate the number of positive labels in the multi-label reference standard. Confidence intervals were computed with 1,000 image-level bootstrap resamples.

| Class | AUROC (95% CI) | AUPRC (95% CI) | Se (95% CI) | Spe (95% CI) | PPV (95% CI) | F1 (95% CI) |
| --- | --- | --- | --- | --- | --- | --- |
| AF (N=7) | 0.996 (0.983-1.000) | 0.948 (0.775-1.000) | 1.000 (1.000-1.000) | 0.979 (0.948-1.000) | 0.778 (0.498-1.000) | 0.875 (0.667-1.000) |
| AT/Flutter (N=10) | 0.935 (0.825-0.998) | 0.786 (0.530-0.982) | 0.600 (0.273-0.909) | 0.989 (0.967-1.000) | 0.857 (0.500-1.000) | 0.706 (0.400-0.933) |
| PSVT/ST (N=6) | 0.985 (0.955-1.000) | 0.850 (0.583-1.000) | 1.000 (1.000-1.000) | 0.928 (0.873-0.979) | 0.462 (0.200-0.750) | 0.632 (0.333-0.857) |
| VT/VF (N=16) | 0.903 (0.791-0.986) | 0.779 (0.564-0.929) | 0.438 (0.167-0.688) | 1.000 (1.000-1.000) | 1.000 (1.000-1.000) | 0.609 (0.286-0.815) |
| AVB 2<sup>nd</sup>/3<sup>rd</sup>/Adv./SB (N=12) | 0.960 (0.909-0.992) | 0.816 (0.611-0.956) | 0.667 (0.375-0.909) | 0.956 (0.907-0.989) | 0.667 (0.375-0.923) | 0.667 (0.400-0.848) |
| ST Elevation (N=0) | — | — | — | 0.990 (0.971-1.000) | 0.000 (0.000-0.000) | — |
| Injury Current (N=35) | 0.988 (0.968-1.000) | 0.971 (0.918-0.999) | 0.686 (0.514-0.833) | 0.985 (0.951-1.000) | 0.960 (0.864-1.000) | 0.800 (0.666-0.900) |
| Premature beat (N=5) | 0.992 (0.970-1.000) | 0.871 (0.452-1.000) | 1.000 (1.000-1.000) | 0.898 (0.827-0.950) | 0.333 (0.091-0.600) | 0.500 (0.182-0.751) |
| RBBB (N=8) | 0.939 (0.824-1.000) | 0.759 (0.409-1.000) | 0.625 (0.250-1.000) | 0.968 (0.929-1.000) | 0.625 (0.222-1.000) | 0.625 (0.249-0.858) |
| LBBB (N=4) | 1.000 (1.000-1.000) | 1.000 (1.000-1.000) | 1.000 (1.000-1.000) | 1.000 (1.000-1.000) | 1.000 (1.000-1.000) | 1.000 (1.000-1.000) |
| PM (N=8) | 0.892 (0.669-1.000) | 0.869 (0.601-1.000) | 0.875 (0.600-1.000) | 0.979 (0.948-1.000) | 0.778 (0.500-1.000) | 0.824 (0.555-1.000) |
| Normal (N=1) | 0.990 (0.970-1.000) | 0.500 (0.250-1.000) | 0.000 (0.000-0.000) | 0.990 (0.970-1.000) | 0.000 (0.000-0.000) | 0.000 (0.000-0.000) |
| Macro mean | 0.962 (0.936-0.983) | 0.832 (0.770-0.917) | 0.717 (0.654-0.778) | 0.972 (0.962-0.981) | 0.622 (0.558-0.686) | 0.658 (0.582-0.712) |

**Supplementary Table 15.** Class-wise AUROC differences between the external Life in the Fast Lane ECG image set and the final InCor-EMG internal test set. Delta values are LITFL minus internal test. This is an unpaired external-domain comparison. Confidence intervals and p values were obtained from 1,000 bootstrap metric draws; p values were adjusted across classes using the Holm method.

| Class | LITFL - Internal $\Delta$ AUROC (95% CI) | Holm-adjusted p |
| --- | --- | --- |
| AF | -0.001 (-0.013 to 0.005) | 1.000 |
| AT/Flutter | -0.042 (-0.147 to 0.023) | 1.000 |
| PSVT/ST | 0.002 (-0.028 to 0.021) | 1.000 |
| VT/VF | -0.096 (-0.207 to -0.013) | 0.060 |
| AVB 2<sup>o</sup>/3<sup>o</sup>/Adv./SB | -0.033 (-0.086 to 0.000) | 0.416 |
| ST Elevation | — | — |
| Injury Current | -0.001 (-0.022 to 0.012) | 1.000 |
| Premature beat | 0.006 (-0.017 to 0.019) | 1.000 |
| RBBB | -0.059 (-0.173 to 0.001) | 0.462 |
| LBBB | 0.015 (0.009–0.022) | 0.000 |
| PM | -0.104 (-0.328 to 0.006) | 1.000 |
| Normal | 0.022 (0.000–0.036) | 0.372 |

## Notes

### Competing Interest Statement

The authors have declared no competing interest.

### Author Declarations

The study was reviewed by the Ethics Committee / Institutional Review Board of Hospital das Clínicas da Faculdade de Medicina da Universidade de São Paulo (HCFMUSP), São Paulo, Brazil. Ethical approval was granted under CAAE 45070821.3.0000.0068. The requirement for informed consent was waived by the ethics committee because this was a retrospective study using de-identified data.

### Summary of Updates

Change author information. ORCID values were changed for authors

